# Risk-Based Prioritization for COVID-19 Vaccination in the Post-Emergency Phase

**DOI:** 10.1101/2025.11.18.25340522

**Authors:** Matan Yechezkel, Jeremy Samuel Faust, Doron Netzer, Talish Razi-Benita, Erez Shmueli, Ronen Arbel, Dan Yamin

**Affiliations:** School of Public Health, University of California, Berkeley, Berkeley, California, USA; Brigham and Women’s Hospital, Mass General Brigham Division of Health Services Research, Department of Emergency Medicine, Harvard Medical School, Boston, Massachusetts, USA; Community Medical Services Division, Clalit Health Services, Tel Aviv, Israel; MIT Media Lab, Cambridge, MA, USA; Maximizing Health Outcomes Research Lab, Sapir College, Sderot, Israel; Department of Management Science and Engineering, Stanford University, Stanford, California, USA; School of Industrial and Intelligent Systems Engineering, Tel Aviv University, Tel Aviv, Israel

## Abstract

**Importance:** As COVID-19 transitions from an acute emergency to an endemic threat, yearly universal boosting policies may not be necessary. Quantifying absolute benefits of differing age– or risk-based vaccination strategies is critical for efficient allocation.

**Objective:** To estimate the absolute benefit of COVID-19 vaccination strategies in the post-emergency phase based on the number needed to vaccinate (NNV) to prevent moderate-to-severe COVID-19 hospitalization, defined per Israeli Ministry of Health criteria (pneumonia with hypoxia or need for respiratory support), across risk strata.

**Design, Setting, and Participants:** This retrospective, population-based cohort study included all individuals aged 12 years or older enrolled in Clalit Health Services, Israel’s largest healthcare organization, between March 1, 2020, and March 1, 2025. Data from the emergency phase (March 1, 2020 to July 31, 2022) were used to empirically estimate the risk of COVID-19 hospitalization across three dimensions: age group, comorbidity category, where each participant was classified into one of 16 groups according to the condition associated with the highest observed risk, and history of all-cause hospitalizations within the past 1 to 3 years. This risk-based classification, derived from the emergency phase, was then applied to the post-emergency phase to identify high-risk groups.

**Main Outcomes and Measures:** NNV to prevent moderate-to-severe COVID-19 hospitalization.

**Results:** The study included 3.6 million individuals with 39,571 COVID-19–related hospitalizations in the emergency phase and 10,141, in the post-emergency—an effective 4.3 annual decline. During the emergency phase, hospitalization risk ranged from 68 per 100,000 among healthy adults aged 12–49 years to 9,999 per 100,000 among those aged 75 years or older with severe chronic illness and recent hospitalization. In the post-emergency phase, risks declined to 5–3,230 per 100,000 within the same groups. NNV ranged from fewer than 100 in high-risk older adults to over 40,000 in younger healthy adults. Using NNV thresholds of 1,000–2,000, 7.2%–10.0% of the population would be prioritized for vaccination, representing 79%–88% of all COVID-19–related hospitalizations.

**Conclusions and Relevance:** COVID-19 continues to pose considerable risk to vulnerable populations. Consideration of risk factors beyond age can inform more efficient vaccine allocation strategies for ongoing booster doses, particularly by accounting for all-cause hospitalizations in the past 1–3 years.

## BACKGROUND

COVID-19 continues to generate substantial morbidity and mortality, particularly among adults at elevated risk for severe outcomes ^1^. However, as the pandemic has evolved into an endemic phase, new evidence-based public health strategies that prioritize those most likely to benefit from routine vaccinations should be considered ^2,3^. Recent commentaries have emphasized the need to evaluate vaccination with the same standards of evidence, transparency, and proportionality as those applied to other preventive interventions ^1^.

The risk of severe COVID-19 is highly heterogeneous. Age remains the strongest predictor, and multiple underlying medical conditions—including chronic cardiopulmonary disease, immunocompromise, metabolic disorders, chronic kidney disease, and obesity —increase the likelihood of hospitalization and death ^4^. Additional strong marker to individual’s general health is recency of any hospitalization. The observation that an individual needed a severe intervention such as hospitalization and the hospitalization procedure itself (surgical operation, invasive, medication) that may requires considerable time for recovery – provide an indication for a temporal elevated risk, and often neglected in policy recommendations. The distribution of these risk factors within a population significantly affects both the relative and absolute benefits of vaccines, and therefore the anticipated public health impact of risk-based vaccination strategies.

A key unresolved question is how to prioritize post-emergency phase COVID-19 vaccination to maximize clinical benefit while minimizing costs and burden. Therefore, we quantified the absolute benefit of COVID-19 boosters across differing age-, underlying medical conditions and recency of hospitalizations for prioritization strategies by estimating the number needed to vaccinate (NNV) to prevent COVID-19 hospitalizations or death.

## METHODS

### Procedures

We analyzed the electronic health records (EHR) of individuals aged 12 and above who were active members of Clalit Health Services (CHS), the largest healthcare organization in Israel, between March 1, 2020, to March 1, 2025 (Appendix, Figure S1). As the largest healthcare provider in Israel, it provides comprehensive medical coverage for approximately 52% of the population (around 5 million members) and two-thirds of individuals aged ≥65 years. Members of CHS are representative of the Israeli population in terms of age, population sectors and socioeconomic status, ensuring inclusion of all major demographic and ethnic groups ^5^. Records are automatically collected and updated daily in the databases of all CHS medical facilities nationwide. We extracted each patient’s socio-demographic and documented of chronic illnesses diagnoses. The following demographic data were extracted for each participant: age, sex, geographical district of the primary healthcare clinic, population sector (General Jewish, Arab, and Ultra-Orthodox Jews), comorbidities, and scores for socio-economic status (SES), which were derived from small statistical areas defined by the Israeli Central Bureau of Statistics and updated by the POINTS Location Intelligence Company to group the entire CHS population into ten categories (1 = lowest, 10 = highest).

### Study design and participants

We conducted a retrospective, population-based analysis using a unique stratification approach to estimate the risk of COVID-19 hospitalization across age, prior hospitalization history, and comorbidities. The study period encompassed emergency and post-emergency phases of the COVID-19 pandemic, defined as March 1, 2020, to July 30, 2022 and August 1, 2022, to March 1, 2025, respectively. To be included in our analysis, individuals needed to be alive and active members of CHS at the start of the study period and remained active until its end or until death, whichever occurred first.

### Outcomes

The primary outcome measured in this study was moderate or severe COVID-19 hospitalization. Moderate or severe COVID-19 hospitalization was defined according to Israeli Ministry of Health criteria as COVID-19 hospitalization with clinical or radiologic evidence of COVID-19 pneumonia accompanied by oxygen saturation <93% on room air, PaO₂/FiO₂ <300, respiratory distress, or need for respiratory support ^6,7^. Mild COVID-19 hospitalizations were excluded because they were often precautionary or not the primary cause of admission during the emergency phase, and therefore did not reflect clinically significant disease severity. We assessed the average yearly incidence rate of the study outcomes stratified by age, hospitalization history and comorbidities. Additionally, we investigated the absolute risk reduction to assess the NNV to prevent one COVID-19 hospitalization, given disease incidence during the study period.

### Data analysis

In line with US Centers for Disease Control & Prevention (CDC) guidance ^4^, we classed comorbidities recorded in the CHS chronic condition registry were grouped into 16 health-related categories as follows: no documented comorbidities, other immune / endocrine disorders, smoking / alcohol / drug use, chronic lung disease, chronic liver disease, neurological disorders, hypertension, obesity, autoimmune / inflammatory disorders, diabetes mellitus, chronic kidney disease, cardiovascular disease, solid malignancy, severe pulmonary or hepatic disease, transplant / hematologic malignancy / end-stage renal disease (ESRD), and other chronic conditions.

Then, we assigned a risk-based severity ranking to the 16 health-related categories according to each category’s observed association with the severity of COVID-19 hospitalization (Appendix, Table S1). The order of severity was determined in a data-driven manner by analyzing the full emergency-period cohort and ranking the categories according to their absolute risk of COVID-19 hospitalization. Each participant was assigned to one health group on a monthly basis. When multiple conditions were present in a given month, the participant was classified into the group associated with the highest risk of severe COVID-19 outcomes. For example, an individual with both obesity and cardiovascular disease was classified into the cardiovascular disease group, reflecting the principle that the highest-risk condition takes precedence in categorization.

From March 1, 2020, to March 1, 2025, each participant was assigned monthly to a subpopulation defined by characteristics associated with the risk of COVID-19 hospitalization. These characteristics included age (12–49, 50–64, 65–74, and ≥75 years), hospitalization history during the preceding three years (none, any hospitalization within the past year, or hospitalization 2–3 years earlier), and one of 16 health-group categories (Appendix, Table S1). To adjust for differences in vaccination coverage, outcome counts were adjusted to reflect a counterfactual scenario in which no individuals were vaccinated using bootstrap method with 1,000 repetitions. Following a recent meta-analysis ^8^, vaccine effectiveness (VE) was assumed to persist for 180 days, with estimates from 30% to 65% (Appendix, Tables S2 and 3). For each period (emergency and post-emergency phases), we calculated the average annual rate of COVID-19 hospitalization per 100,000 individuals within each subpopulation. Ninety-five percent confidence intervals (CIs) were computed assuming a binomial distribution, with the total number of individuals in each subpopulation representing the number of trials and the proportion hospitalized representing the probability of the event. We then estimated the NNV with the COVID-19 vaccine to prevent one hospitalization across all subpopulations. NNV was derived from the absolute risk reduction.

### Sensitivity analysis

As a sensitivity analysis, we estimated the share of COVID-19 hospitalizations that would be covered under alternative vaccination strategies, in which vaccination was offered to all individuals with a NNV ≤ 500, 1000, or 2000. Coverage was evaluated across VE values ranging from 30% to 65%, weighting by population size. Ninety-five percent confidence intervals were derived using a multinomial bootstrap of subgroup-specific hospitalization counts, accounting for sampling variability while keeping subgroup membership fixed.

We also calculated for different NNV cutoffs ranging from 500-2000, the proportion of the general population that would need to be vaccinated. CHS accounts for 52% of the population ^5^, but for a better representation of the total population, we used data on age distribution from the Central Bureau of Statistics and adjusted the required vaccination coverage to the general population^9^.

### Post-hoc analysis

To determine the independent and joint associations of age, health-related category, and prior hospitalization with the risk of COVID-19 hospitalization, we used logistic regression with all main effects, two-way interactions, and a three-way interaction term (*Age × Health group × Hospitalization history*). P-values were derived from Wald tests to assess statistical significance.

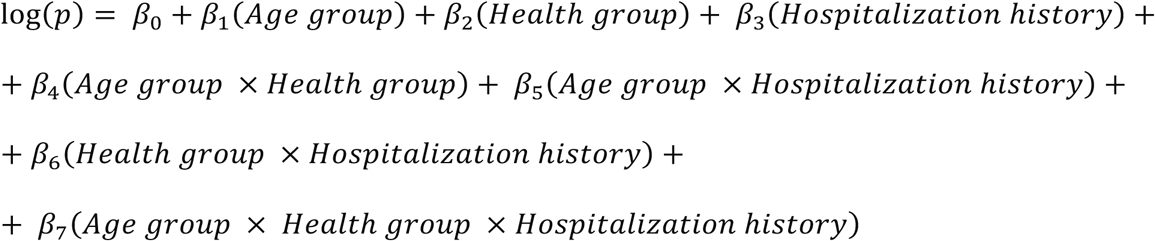

#### Ethical Approval

As the retrospective data were pseudonymized, the Clalit Health Services institutional Helsinki review board and data utilization committee approved the use of the retrospective cohort data without requiring specific consent from the members of Clalit Health Services (protocol number 0139-21-CHS).

## RESULTS

### Study population

The cohort included 3,633,478 CHS members. Of these, 3,633,478 and 3,535,201 were represented in the emergency and post-emergency phases, respectively. The mean ages were similar, 42 and 44 years, respectively. 925,511 (25.5%) in the emergency period and 797,799 (22.6%) in the post-emergency had no documented comorbidities (Table 1). During the emergency, 2,656,682 (73.1%) received two doses of a COVID-19 vaccine, and 2,036,534 (56.0%) received at least one booster dose. In contrast, during the post-emergency phase, vaccination coverage was markedly lower, with 121,563 (3.4%) of the cohort vaccinated with a booster. During the emergency phase, 39,571 moderate-to-severe COVID-19 hospitalizations were observed, declining to 10,141 in the post-emergency phase.

**Table 1.** Cohort characteristics.

| Characteristic | Emergency (n=3,633,478) | Post-emergency (n=3,535,201) |
| --- | --- | --- |
| **COVID-19 hospitalizations** | 39,571 | 10,141 |
| **Sex** |  |  |
| Female | 51.5% (1,869,543) | 51.5% (1,821,208) |
| Male | 48.5% (1,763,935) | 48.5% (1,713,993) |
| **Age group [years]** |  |  |
| Median (IQR) | 42 (28-62) | 44 (30-63) |
| 12-49 | 61.5% (2,234,887) | 58.8% (2,078,011) |
| 50-64 | 16.6% (603,898) | 17.8% (628,759) |
| 65-74 | 10.9% (397,153) | 11.2% (394,856) |
| 75+ | 10.9% (397,540) | 12.3% (433,575) |
| **Sociodemographic score** |  |  |
| 1-4 | 34.5% (1,254,087) | 34.6% (1,224,344) |
| 5-7 | 40.4% (1,467,956) | 40.2% (1,422,749) |
| 8-10 | 19.9% (722,961) | 19.9% (702,868) |
| Unspecified | 0.2% (5,986) | 0.2% (5,698) |
| **Health group** |  |  |
| Transplant / Hematologic Malignancy / ESRD | 1.6% (58,830) | 1.7% (61,690) |
| Severe Pulmonary or Hepatic Disease | 3.2% (116,657) | 3.3% (116,402) |
| Solid Malignancy | 5.1% (185,809) | 5.5% (195,956) |
| Cardiovascular Disease | 25.9% (942,094) | 27.0% (954,218) |
| Chronic Kidney Disease | 2.0% (71,271) | 2.2% (76,463) |
| Diabetes Mellitus | 0.8% (29,684) | 0.8% (27,696) |
| Autoimmune / Inflammatory Disorders | 7.0% (254,622) | 7.3% (257,277) |
| Obesity | 7.2% (262,498) | 7.9% (277,589) |
| Hypertension | 0.7% (26,073) | 0.7% (24,673) |
| Neurological Disorders | 0.9% (31,529) | 0.9% (31,901) |
| Chronic Liver Disease | 0.3% (12,692) | 0.4% (12,509) |
| Chronic Lung Disease | 3.3% (121,282) | 3.3% (118,320) |
| Other Conditions | 2.8% (101,609) | 3.0% (105,134) |
| Smoking / Alcohol / Drug Use | 9.7% (351,136) | 9.2% (326,379) |
| Other Immune / Endocrine Disorders | 3.9% (142,181) | 4.3% (151,195) |
| No Documented Comorbidities | 25.5% (925,511) | 22.6% (797,799) |
| **Population sector** |  |  |
| General Jewish | 67.5% (2,453,694) | 67.6% (2,388,189) |
| Ultra-Orthodox Jewish | 5.6% (204,568) | 5.7% (201,499) |
| Arab | 25.4% (921,260) | 25.8% (910,368) |
| Other | 1.5% (53,956) | 1.0% (35,145) |
| **Vaccination status** |  |  |
| Unvaccinated | 26.9% (976,796) | 25.8% (913,623) |
| Primary series | 17.1% (620,148) | 17.1% (604,419) |
| First booster | 44.7% (1,622,918) | 45.5% (1,609,442) |
| Second booster | 11.3% (413,616) | 8.1% (286,154) |
| Third booster | NA | 3.4% (121,563) |

### Absolute Risk for Severe Covid-19 Outcomes

During the emergency period, the risk of COVID-19–related moderate-to-severe hospitalization varied markedly across age and health groups (Figure 1; Appendix Table S2). Among the lowest-risk groups, no documented comorbidities individuals aged 49 years or younger with no hospitalizations in the preceding 3 years had an average annual hospitalization rate of 68 per 100,000 (95% CI, 64 to 71). In contrast, adults aged 75 years or older with a history of transplant and a hospitalization in the preceding year had an exceptionally high rate of 9,999 per 100,000 (95% CI, 9,441 to 10,581). Substantial heterogeneity was also observed within the oldest age group: among individuals 75 years or older, the rate ranged from 125 per 100,000 (95% CI, 88 to 172) among no documented comorbidities adults with no recent hospitalization to 9,999 per 100,000 (95% CI, 9,441 to 10,581) among those with transplant, hematologic malignancy, or end-stage renal disease and a recent hospitalization (Figure 1).

**Figure 1.**
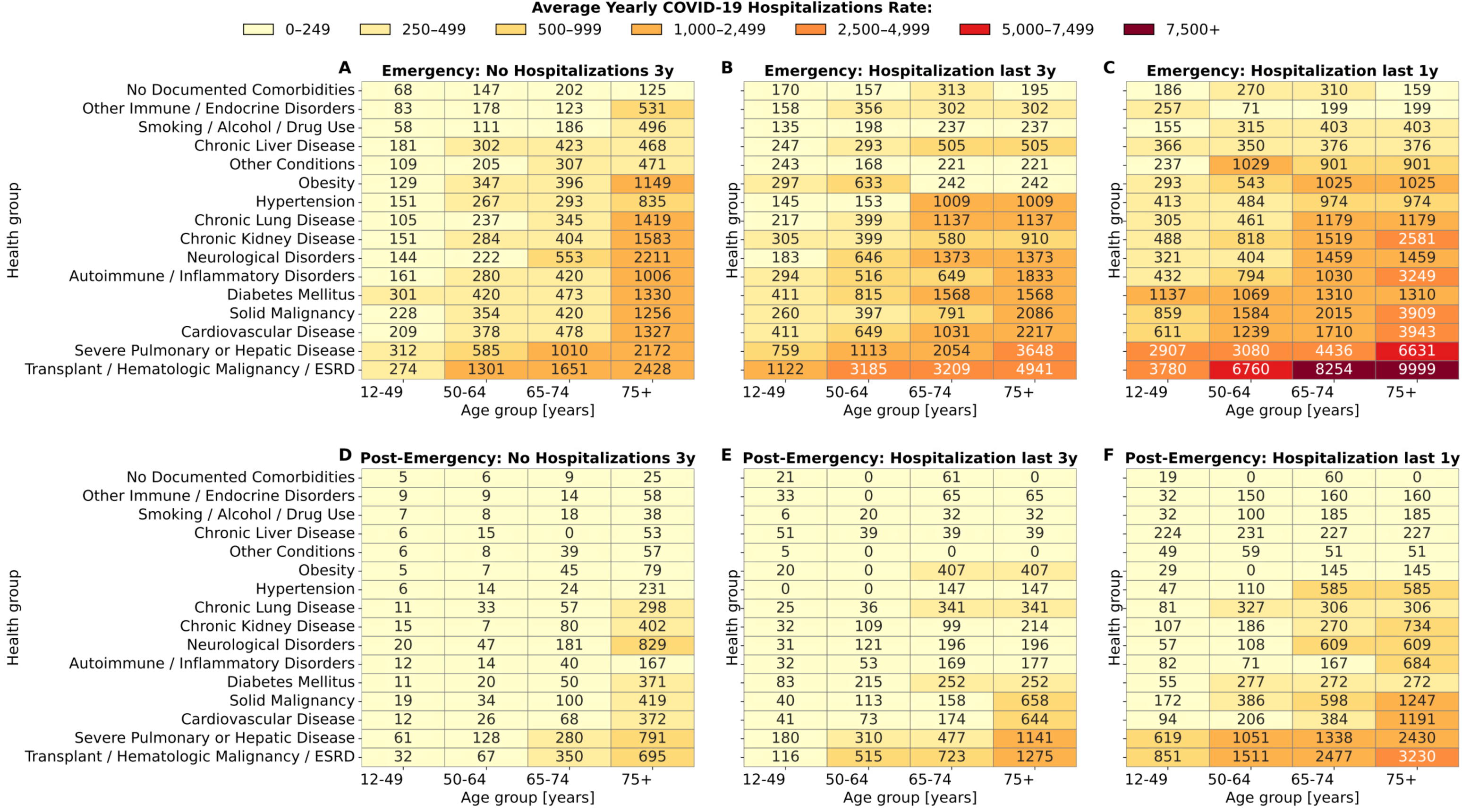
Impact of age, comorbidities, and hospitalization history on covid-19 hospitalization rates. Average annual rates of COVID-19–related hospitalizations per 100,000 individuals, stratified by age group, comorbidity category, and history of prior hospitalizations, are observed for the emergency (A–C) and post–emergency (D–F) periods. To avoid sparse cells, strata with <500 individuals were pooled across adjacent age groups, keeping health-group and hospitalization-history unchanged. 95% CIs are reported in Appendix Tables S2 and 3. Definitions of health groups are detailed in Appendix Table S1.

The logistic regression analysis showed a significant association between prior hospitalization and subsequent COVID-19 hospitalization (p < 0.001), with evidence of interaction across age and health-related categories. Individuals hospitalized in the prior year were approximately 7.7 times more likely to be hospitalized with COVID-19, and those hospitalized 1–3 years earlier were 3.4 times more likely, consistent with the descriptive analysis (Appendix Table S5).

Variation in risk across population strata persisted in the post-emergency phase, despite a reduction in absolute risk across all groups (Figure 1; Appendix Table S3). For example, among adults aged 75 years or older with a recent hospitalization and a history of transplant, the hospitalization rate was 3,230 per 100,000 (95% CI, 2,951 to 3,528), compared with 25 per 100,000 (95% CI, 8 to 59) among individuals of the same age without documented comorbidities or recent hospitalization (Figure 1). Moreover, recent hospitalization was a pivotal indicator of elevated risk: compared with those not hospitalized in the prior three years, individuals with a hospitalization in the previous year were 13.2 times more likely to be hospitalized with COVID-19, while those hospitalized 1–3 years earlier were 4.5 times more likely.

### Number Needed to Vaccinate to Prevent Covid-19 Hospitalizations in the post emergency era

The NNV to prevent 1 moderate-to-severe COVID-19 hospitalization varied substantially across age groups and comorbidity categories, with recency of hospitalization emerging as a particularly strong determinant (Figure 2; Appendix Table S4). Among adults aged 75 years or older with severe pulmonary or hepatic disease, the NNV was 91 (95% CI, 64-133) when a hospitalization had occurred in the preceding year, compared with 280 (95CI, 197-410) when no hospitalization had occurred in the prior 3 years. Similarly, among adults aged 65 to 74 years with chronic lung disease, the NNV was 708 (95% CI, 528-1058) with a hospitalization in the preceding year versus 3,891 (95% CI, 2,734-5,689) with no hospitalization in the prior 3 years.

**Figure 2.**
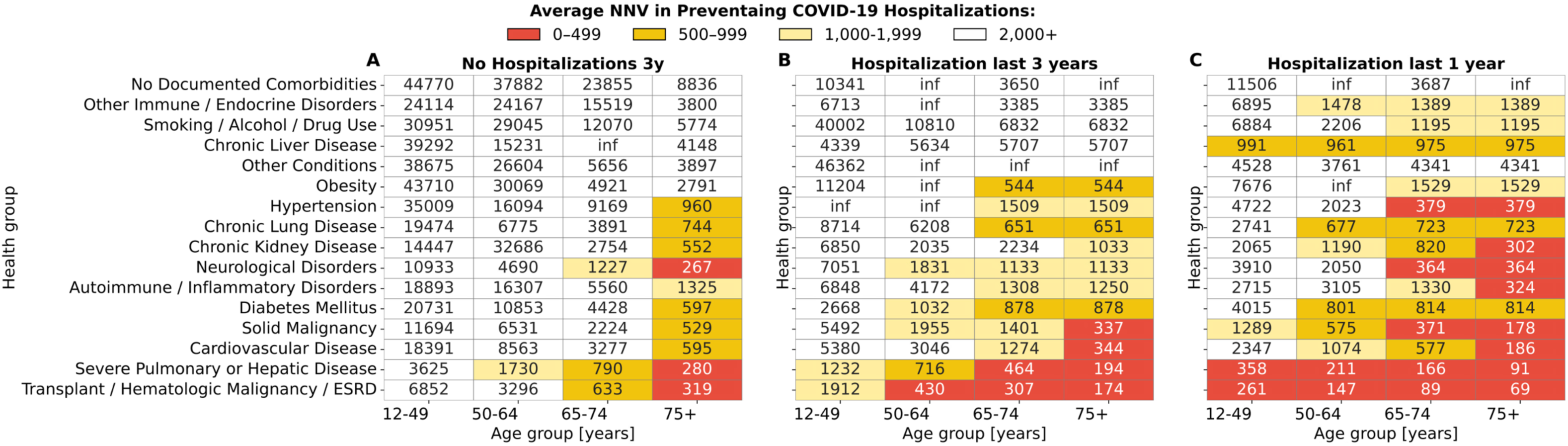
Impact of age, comorbidities, and hospitalization history on number needed to vaccinate to prevent one covid-19 hospitalization during the post-emergency period. 95% CIs are provided in Appendix, Table S4. Definitions of health groups are detailed in Appendix Table S1.

Considering an NNV cutoffs of 1,000 and 2,000 to guide vaccination prioritization, individuals with chronic lung disease should be targeted only if they are aged 65 years or older and had a non–COVID-19 hospitalization in the preceding year (Table 2). A similar pattern was observed for individuals whose most severe comorbidity was hypertension. In contrast, adults aged 75 years or older with cardiovascular disease should be prioritized regardless of hospitalization history, with the age threshold lowered to 50 years or older for those hospitalized in the prior year.

**Table 2.** Identification of individuals for vaccination prioritization using an NNV cutoff of 1,000 and 2,000.

| Health group / NNV cutoff | <u>No Hospitalizations in the</u><br><u>last 3 years</u> |  | <u>Hospitalization in the</u><br><u>prior 2-3 years</u> |  | <u>Hospitalization in the</u><br><u>last 1 year</u> |  |
| --- | --- | --- | --- | --- | --- | --- |
|  | 1,000 | 2,000 | 1,000 | 2,000 | 1,000 | 2,000 |
| No Documented Comorbidities | — | — | — | — | — | — |
| Other Immune / Endocrine Disorders | — | — | — | — | — | 50+ |
| Smoking / Alcohol / Drug Use | — | — | — | — | — | 65+ |
| Chronic Liver Disease | — | — | — | — | 12+ | 12+ |
| Other Conditions | — | — | — | — | — | — |
| Obesity | — | — | 65+ | 65+ | 65+ | 65+ |
| Hypertension | 75+ | 75+ | 75+ | 65+ | 65+ | 65+ |
| Chronic Lung Disease | 75+ | 75+ | 65+ | 65+ | 50+ | 50+ |
| Chronic Kidney Disease | 75+ | 75+ | 75+ | 75+ | 65+ | 50+ |
| Neurological Disorders | 75+ | 65+ | 75+ | 50+ | 65+ | 50+ |
| Autoimmune / Inflammatory Disorders | — | 75+ | — | 65+ | 75+ | 65+ |
| Diabetes Mellitus | 75+ | 75+ | 65+ | 50+ | 50+ | 50+ |
| Solid Malignancy | 75+ | 75+ | 75+ | 50+ | 50+ | 12+ |
| Cardiovascular Disease | 75+ | 75+ | 75+ | 65+ | 65+ | 50+ |
| Severe Pulmonary or Hepatic Disease | 65+ | 50+ | 50+ | 12+ | 12+ | 12+ |
| Transplant / Hematologic Malignancy / ESRD | 65+ | 65+ | 50+ | 12+ | 12+ | 12+ |

Based on NNV values appeared in Figure 2 and Appendix Table S4.

### Sensitivity Analysis

The sensitivity analysis indicated that vaccination prioritization and coverage would not change substantially when applying different NNV thresholds between 1,000 and 2,000 (Figure 3). Specifically, using the cutoff of 1,000, 7.2% (95% CI, 6.7-9.4) of the population in the post–COVID-19 emergency period would be prioritized for vaccination, compared with 10.0% (95% CI, 9.2-12.4) when applying a cutoff as high as 2,000 (Figure 3). Applying an NNV threshold of 1,000 was estimated to cover 79%–85% of COVID-19 hospitalizations across VE values from 30% to 65%, compared with 85%–88% when using a threshold of 2,000 (Figure 4).

**Figure 3.**
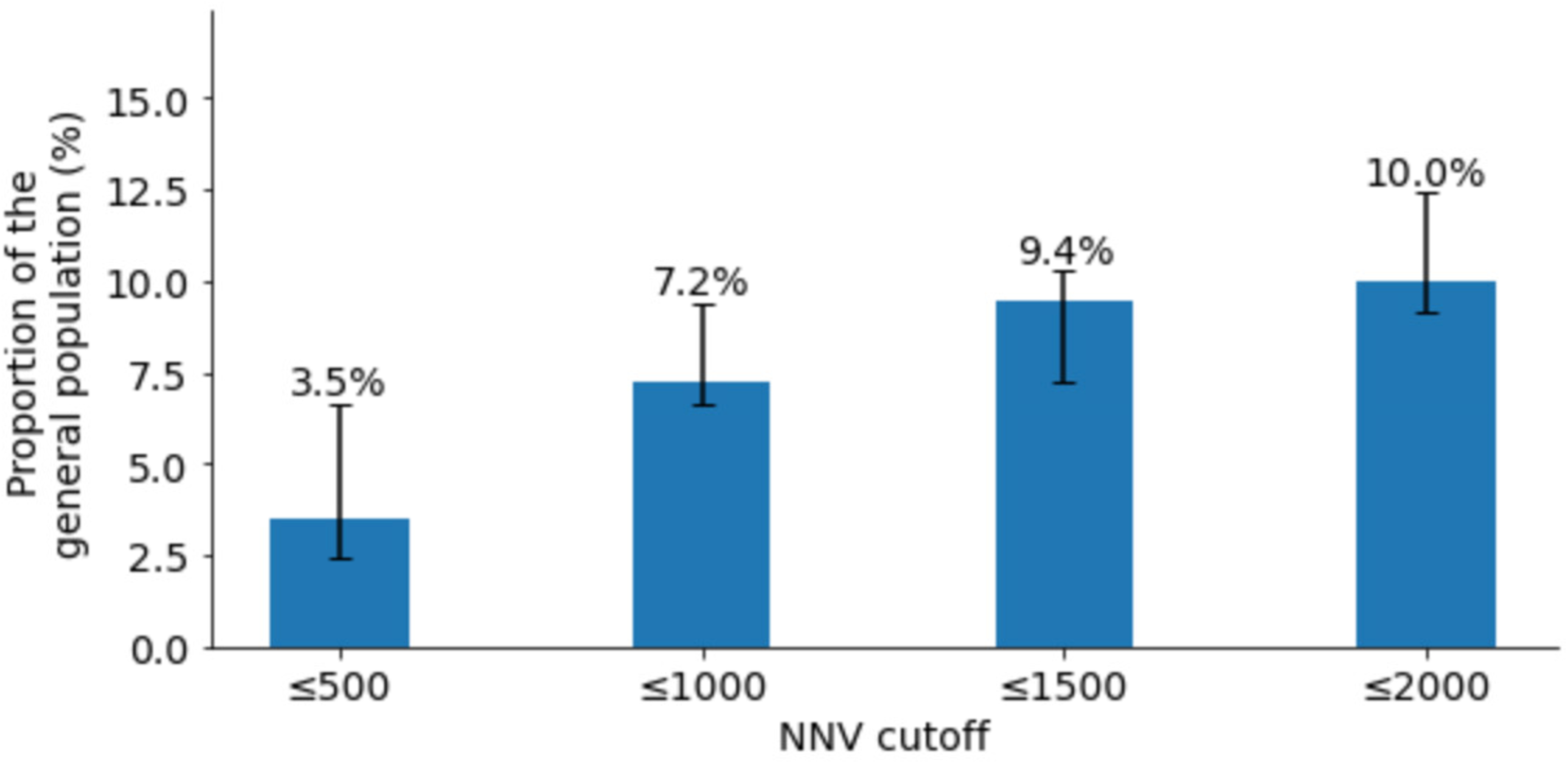
Estimated proportion of the general population requiring prioritization for COVID-19 vaccination under varying NNV thresholds during the post-emergency phase. 95% CIs were derived assuming binomial distribution for COVID-19 hospitalization risks.

**Figure 4.**
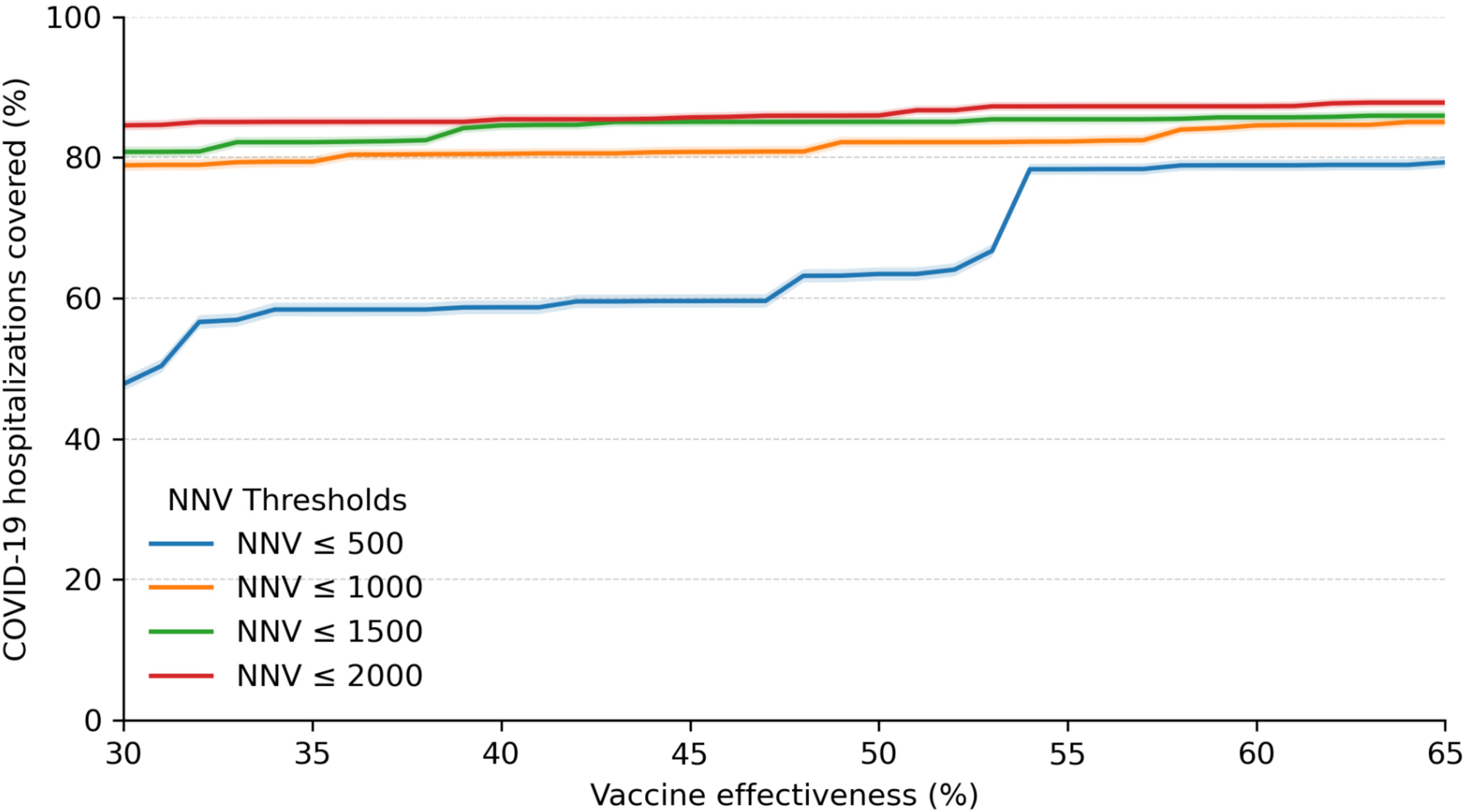
Estimated proportion of COVID-19 hospitalization covered under different NNV cutoffs and VE values of 30-65% during the post-emergency phase. 95% CIs were calculated using percentile bootstrap method with 1,000 repetitions.

## DISCUSSION

### Summary of Results

In this real-world, nationwide analysis from CHS, the risk of moderate-to-severe COVID-19 hospitalization during the post-emergency period varied by orders of magnitude across strata defined by age, comorbidity severity, and recent non–COVID–19 hospitalization. Absolute risks were, as expected, lower than during the emergency period but remained concentrated among the oldest adults and those with severe underlying conditions or recent hospitalizations. Translating these risks into absolute benefits, the NNV to avert one hospitalization differed sharply across strata, yielding small NNVs (indicating a greater absolute benefit) among the very old and immunocompromised, and very large NNVs among younger adults without major comorbidities.

As Most studies evaluating the effectiveness of COVID-19 vaccines report Relative Vaccine Efficacy (RVE), a statistical measure that compares outcomes between vaccinated individuals and an unvaccinated control group. While RVE is useful for demonstrating comparative benefit, it does not provide actionable insight into the absolute impact of vaccination, particularly regarding the number of individuals who need to be vaccinated to prevent severe outcomes such as hospitalization.

Our article introduces and emphasizes the NNV as a critical metric for health policy and resource allocation. NNV quantifies the number of people who must receive the vaccine to prevent one hospitalization, offering a practical and transparent measure of real-world benefit. By applying NNV, decision-makers in health organizations can better identify and prioritize high-risk populations, ensuring that vaccination strategies are both clinically effective and economically efficient. Our analysis underscores the importance of shifting from relative measures to absolute metrics in guiding public health interventions and optimizing vaccine deployment.

### Comparison to Current Evidence

Our findings align with contemporary evidence that VE in 2024–2025 remains clinically meaningful against medically attended illness and severe outcomes, while showing heterogeneity by clinical setting and time since vaccination ^10^. By expressing benefits as NNV across granular risk strata, our results operationalize the evidence-based prioritization principles recently emphasized for post-emergency vaccination policy, including attention to absolute risk, transparency, and proportionality ^1^. They also align with prior analyses, which demonstrate that updated boosters reduced severe outcomes during Omicron waves, with effect sizes being largest in the oldest and most clinically vulnerable groups ^4^. Finally, our risk-gradient results mirror established epidemiology, which indicates that severe COVID-19 outcomes cluster in individuals with advanced age and specific chronic conditions (e.g., immunocompromise, chronic lung or kidney disease, metabolic disorders), supporting targeted approaches consistent with clinical guidance.

### Limitations

This observational study is subject to residual confounding and misclassification. Although we stratified the data finely by age, comorbidity, and recent non–COVID–19 hospitalization, unmeasured factors (e.g., functional status, frailty indices, household composition) may still influence both vaccination and outcomes. Second, hospitalization outcomes and comorbidity assignments rely on administrative data and may be affected by changes in testing practices, admission thresholds, and coding over time. Third, VE assumptions used to translate absolute risks into NNV were based on contemporary external estimates and scenario ranges ^10^; For example, if actual VE differs materially by subpopulation or circulating lineage, NNV estimates would shift accordingly. Fourth, our data arises from a single, integrated health system within Israel. At the same time, CHS members are socio-demographically diverse, and the generalizability of our findings to other countries and healthcare contexts may be imperfect. Fifth, our analysis focuses on the prevention of hospitalization; it does not quantify the vaccine’s impact on long COVID, transmission, or healthcare workforce absenteeism, nor does it address vaccine safety or formal cost-effectiveness—issues relevant to comprehensive policy decisions. A limitation of this analysis is that vaccine effectiveness may vary over time, and the estimated NNVs could change depending on the timing of vaccination relative to infection waves. In some subgroups, such as older adults with diabetes, the association between prior hospitalization and subsequent COVID-19 hospitalization did not follow the expected pattern, which may reflect differences in comorbidity profiles or reasons for previous admissions. Finally, the choice of any NNV cutoff is ultimately normative and context dependent (budget, logistics, equity goals) and should be revisited as epidemiology and program capacity change.

### Implications for Policy

First, booster policies should explicitly anchor on absolute risk reduction, which is quantified by NNV. Health systems can implement stratified invitations keyed to age bands, comorbidity class, and recent hospitalization—variables that are routinely available in electronic health records—to identify those with the smallest NNVs. Second, the program scope may justifiably contract in the post-emergency period: using an NNV cutoff such as 1,000 –2000 ^11^, only a modest share of the population would be prioritized at any given time, concentrating resources where the absolute benefit is greatest, focusing vaccination programs on the 7-10% of the population that accounts for the 79-88% of preventable hospitalizations, while substantially reducing the programmatic scope compared with emergency-era universal strategies.

Vaccination coverage among high-risk groups remains markedly low in the post-emergency era despite strong evidence that booster doses are safe and effective^8,12–15^. Transparent communication about absolute risk and NNV can improve public trust and decision quality and provide a consistent framework for adjusting prioritization as variants, VE, and population immunity evolve.

## Conclusions

In the current post-emergency COVID-19 era, one-size-fits-all booster strategies are unlikely to be optimal. A risk-based approach that targets older adults and those with severe underlying conditions or recent hospitalizations yields the greatest absolute benefit per dose and can substantially reduce program scope without compromising the prevention of severe outcomes. Our results provide an operational blueprint—grounded in routine electronic records—for prioritizing COVID-19 vaccination using absolute risk and NNV. Health systems and policymakers can use this framework to calibrate cadence and eligibility as variants, VE, and population immunity evolve, while maintaining equity by proactively reaching high-risk groups most likely to benefit.

## Data Availability

According to this study's CHS Helsinki and data utilization committees' guidelines, no patient-level data is to be shared outside the permitted researchers.

## Acknowledgement

This research was partially supported by the European Research Council (ERC) project #949850 and the Israel Science Foundation (ISF), grant No. 3409/19, within the Israel Precision Medicine Partnership program.

## Authors’ contributions

Conception and design: DY, MY, ES, RA, and JSF. Collection and assembly of data: TR and MY, Analysis and interpretation of the data: MY, DY, ES, RA, and JSF. Statistical expertise: MY, DY, ES. Drafting the article: MY, RA, and DY. Critical revision of the article for important intellectual content: JSF, MY, DY, ES, RA, and DN. Final approval of the article: All authors. Obtaining funding: DY, ES.

## Supplementary Materials for

## Appendix A – Study protocol

### Description of the data

Data will be extracted from the Clalit Healthcare Services (CHS) database. As the largest nationwide health plan (payer–provider) in Israel, it provides comprehensive medical coverage for approximately half of the population. The CHS database contains longitudinal data on a stable population of 4.7 million people since 1993 (with <1%/year moving out) ^1^. Data are automatically collected and includes comprehensive laboratory data from a single central lab, full pharmacy prescription and purchase data, and extensive demographic data on each patient. Within this database, diagnoses are recorded using the International Classification of Diseases, Ninth Revision, Clinical Modification (ICD-9-CM) system, supplemented by internally developed coding schemes that allow more granular diagnostic information beyond the ICD codes. Medications are coded according to the Israeli coding system with translations to anatomical therapeutic chemical (ATC) classification system wherever available. Procedures are coded using Current Procedural Terminology (CPT) codes. We will access to the following data for each patient:

➢ Socio-demographics
  - Sex (binary)
  - Age (year of birth)
  - Socioeconomic status by address and according clinic when address is missing) (scale 1-10)
  - Sector (clinic level data – Arab / Jewish/ Ultra-Orthodox Jewish)
  - Nursing home residency (binary)
➢ Comorbidities
  - Chronic diseases (binary classifcation)
    ○ History of malignancies and active malignancies
    ○ Dementia
    ○ Cardiovascular diseases (ischaemic heart disease, cerebrovascular disease/ all cardio sub registries).
    ○ Diabetes (taken from CRI registry)
    ○ Hypertension (taken from CRI registry)
    ○ Asthma
    ○ Chronic Lung Disease
    ○ Rheumatologic diseases
    ○ Chronic Kidney Disease
    ○ Immunocompromised Status
    ○ Chronic Liver Disease
    ○ Organ transplantation
    ○ Additional Chronic Diseases
➢ Vaccination records
➢ Laboratory test results ( binary classification for existence of infectious diseases)
➢ Hospitalization history ({ admission data, primary service, duration, ICD diagnosis code })
➢ Outpatient history (admission data, primary service, ICD diagnosis code)
➢ BMI ({date, value}}
➢ Smoking status {date, yes/no}

### Explanation on the Socio-Economic Status (SES)

Measure SES was based on the small statistical areas (SSA) used in the 2008 Israeli census ^2^. The SSAs contain 3000–4000 people and are created to maintain homogeneity in terms of the sociodemographic composition of the population. The Israeli Central Bureau of Statistics (CBS) utilized demography, education, employment, housing conditions, and income to define the SSAs, and these were grouped into 20 categories ^3^. This data was updated by the POINTS Location Intelligence Company to improve the accuracy of the SES measure, using up-to date sociodemographic, commercial, and housing data. The entire CHS population was grouped into ten categories, ranging from 1 (lowest) to 10 (highest).

### Data collection and storage

We will receive access to the data from the medical records of all members of CHS aged 12 years and older. As the largest nationwide health plan (payer–provider) in Israel, it provides comprehensive medical coverage for approximately half of the population. The CHS database contains longitudinal data on a stable population of 4.7 million people since 1993 (with <1%/year moving out) ^1^. Data are automatically collected and includes comprehensive laboratory data from a single central lab, full pharmacy prescription and purchase data, and extensive demographic data on each patient. Within this database, diagnoses are recorded using the International Classification of Diseases, Ninth Revision, Clinical Modification (ICD-9-CM) system, supplemented by internally developed coding schemes that allow more granular diagnostic information beyond the ICD codes. Medications are coded according to the Israeli coding system with translations to anatomical therapeutic chemical (ATC) classification system wherever available. Procedures are coded using Current Procedural Terminology (CPT) codes. As for the medical data, we will receive access to the electronic health records (EHR) data after the following pseudonymisation procedures:

**1.** Healthcare identification number of the members will be coded.
**2.** Only year of birth is provided.
**3.** Free text is removed. This means any text that was types/recorded/scanned manually by healthcare staff and is not structured in the electronic system. This includes any documented conversations between healthcare staff and patient or summary of from meetings.
**4.** No audio, photos including scanned text, or video contents are provided.

The data access will be conducted using the CHS remote environment. The data are coded, viewed, stored and process only within the Clalit research environment. The researchers will connect to the research environment via Horizon client platform, which is approved by the Ministry of Health. The user connects through a secure connection using Lightweight Directory Access Protocol and two factor authentication system.

### Procedures

We will analyze the EHRs of individuals aged 12 and above who are members of the largest healthcare organization in Israel, CHS, from March 1, 2020, to March 1, 2025. As the largest healthcare provider in Israel, it provides comprehensive medical coverage for approximately 52% of the population (around 5 million members) and two-thirds of individuals aged ≥65 years. Members of CHS are representative of the Israeli population in terms of age, population sectors and socioeconomic status, ensuring inclusion of all major demographic and ethnic groups ^1^. Records are automatically collected and updated daily in the databases of all CHS medical facilities nationwide. The study data are coded, pseudonymized, stored, and processed within the CHS research environment. We extracted each patient’s socio-demographic and potential chronic illnesses information. The following demographic data will be extracted for each participant: age, sex, geographical district of primary healthcare clinic, population sector (General Jewish, Arab, and Ultra-orthodox Jews), and the score for socio-economic status (SES), types of co-morbidities group (see Table S1).

**Table S1.**
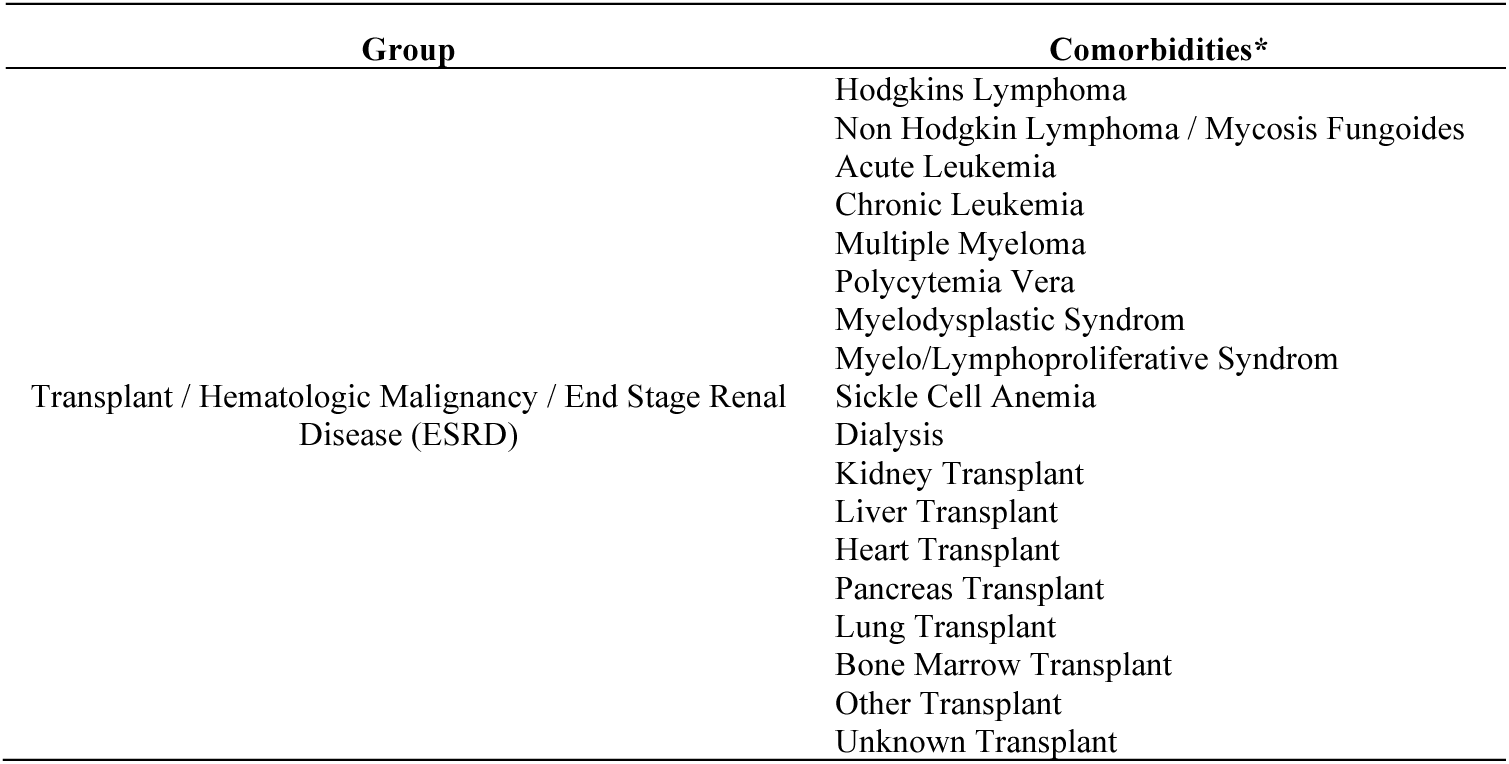
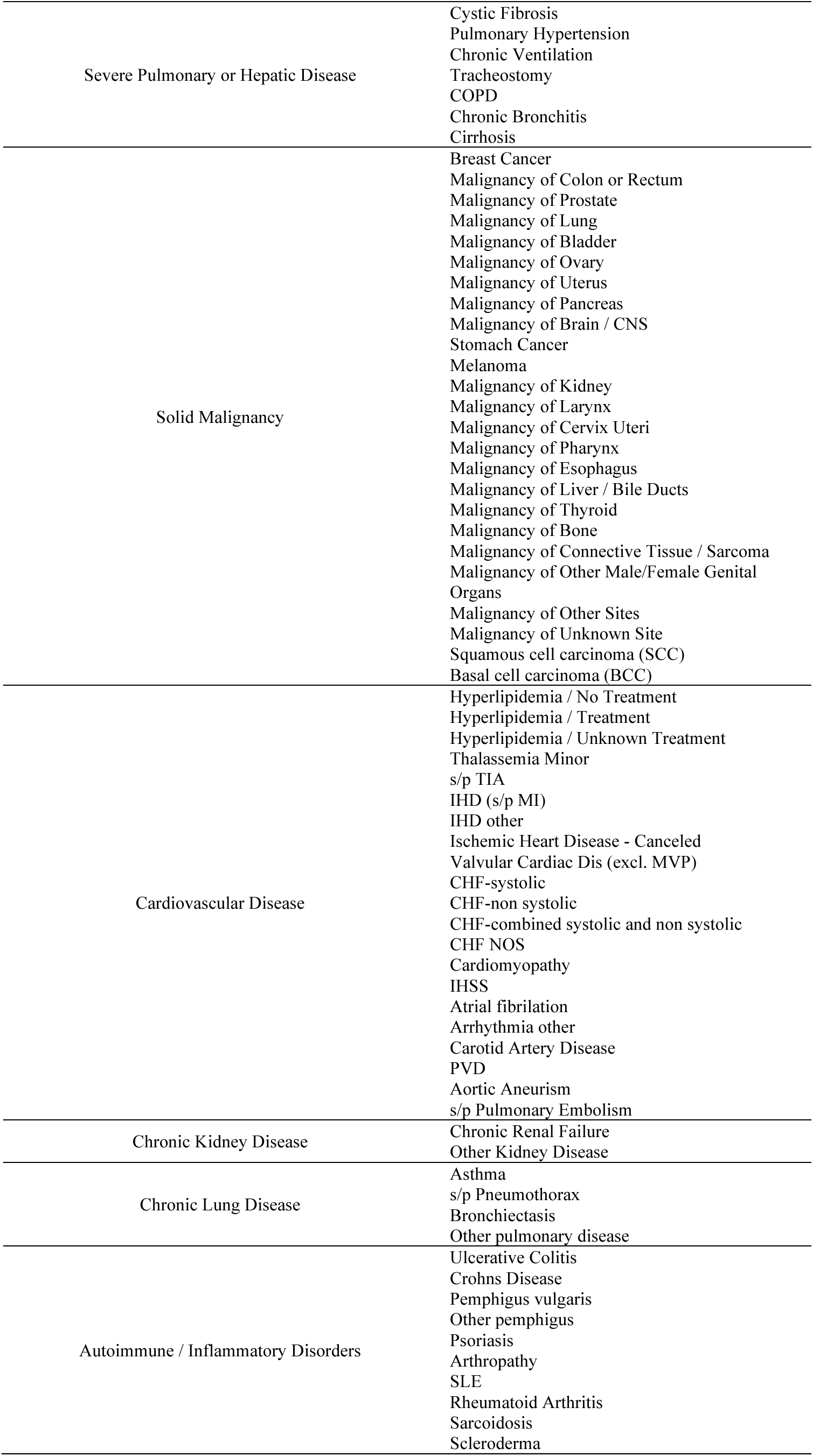
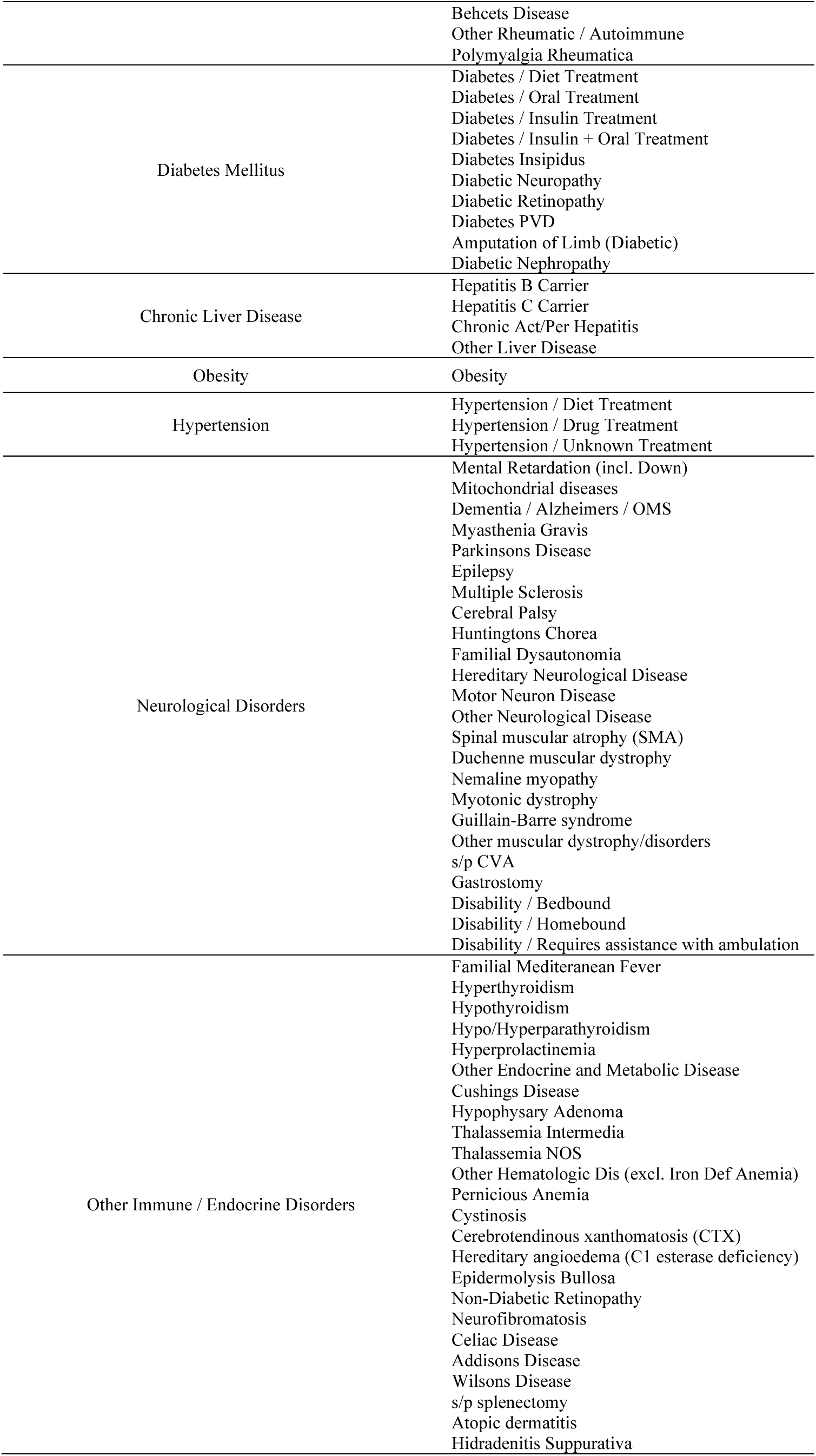
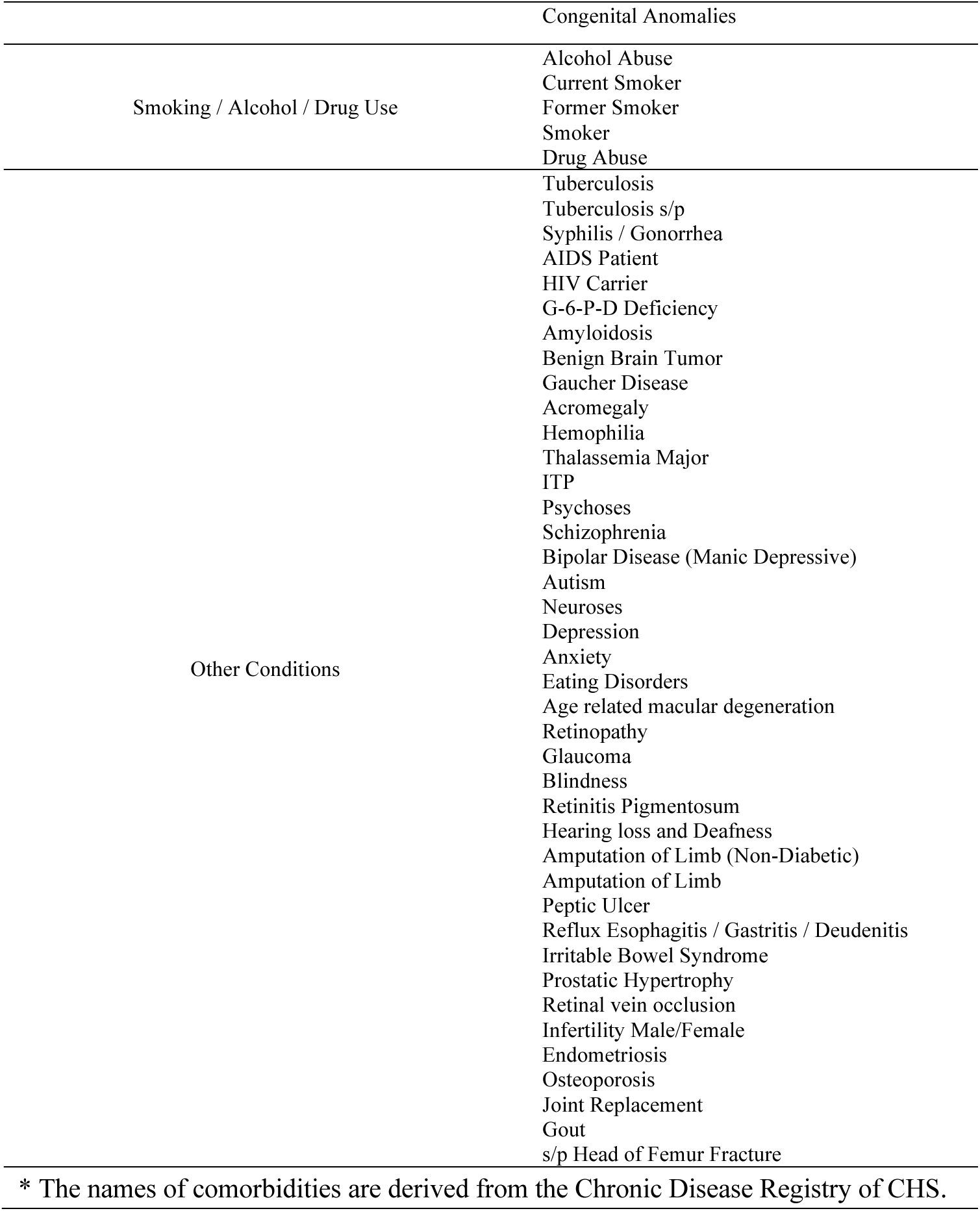
Comorbidity groups.

### Study design and participants

We will conduct a retrospective, population-based analysis using a unique stratification approach to estimate the risk of COVID-19 hospitalization across age, prior hospitalization history, and comorbidities. The study period will encompass emergency and post-emergency phases of the COVID-19 pandemic, defined as March 1, 2020, to July 30, 2022, and August 1, 2022, to March 1, 2025, respectively. To be included in our analysis, individuals need to be alive and active members of CHS at the start of the study period and remain active until its end or until death, whichever occurred first.

### Outcomes

The primary outcome measured in this study is moderate or severe COVID-19 hospitalization. Moderate or severe COVID-19 hospitalization is defined according to Israeli Ministry of Health criteria as COVID-19 hospitalization with clinical or radiologic evidence of COVID-19 pneumonia accompanied by oxygen saturation <93% on room air, PaO₂/FiO₂ <300, respiratory distress, or need for respiratory support ^4,5^. We will exclude mild COVID-19 hospitalizations because they were often precautionary or not the primary cause of admission during the emergency phase and therefore did not reflect clinically significant disease severity. We will assess the average yearly incidence rate of the study outcomes stratified by age, hospitalization history and comorbidities. Additionally, we will investigate the absolute risk reduction to assess the number needed to vaccinate (NNV) to prevent one COVID-19 hospitalization, given disease incidence during the study period.

### Statistical analysis

In line with US Centers for Disease Control & Prevention (CDC) guidance, we will class comorbidities recorded in the CHS chronic condition registry were grouped into 16 health-related categories as follows: no documented comorbidities, other immune / endocrine disorders, smoking / alcohol / drug use, chronic lung disease, chronic liver disease, neurological disorders, hypertension, obesity, autoimmune / inflammatory disorders, diabetes mellitus, chronic kidney disease, cardiovascular disease, solid malignancy, severe pulmonary or hepatic disease, transplant / hematologic malignancy / end-stage renal disease (ESRD), and other chronic conditions (Table S1)^6^.

Then, we will compute a risk-based severity ranking to the 16 health-related categories according to each category’s observed association with the severity of COVID-19 hospitalization. The order of severity will be determined in a data-driven manner by analyzing the full emergency-period cohort and ranking the categories according to their absolute risk of COVID-19 hospitalization. Each participant will be assigned to one health group on a monthly basis. When multiple conditions are present in a given month, the participant will be classified into the group associated with the highest risk of severe COVID-19 outcomes.

From March 1, 2020, to March 1, 2025, each participant will be assigned monthly to a subpopulation defined by characteristics associated with the risk of COVID-19 hospitalization. These characteristics will include age (12–49, 50–64, 65–74, and ≥75 years), hospitalization history during the preceding three years (none, any hospitalization within the past year, or hospitalization 2–3 years earlier), and one of 16 health-group categories (Appendix, Table S1). To adjust for differences in vaccination coverage, outcome counts will be adjusted to reflect a counterfactual scenario in which no individuals were vaccinated using bootstrap method with 1,000 repetitions. Following a recent meta-analysis ^7^, vaccine effectiveness (VE) will be assumed to persist for 180 days, with estimates values from 30% to 65% (Tables S2 and 3). For each period (emergency and post-emergency phases), we will calculate the average annual rate of COVID-19 hospitalization per 100,000 individuals within each subpopulation. Ninety-five percent confidence intervals (CIs) will be computed assuming a binomial distribution, with the total number of individuals in each subpopulation representing the number of trials and the proportion hospitalized representing the probability of the event. We then will estimate the NNV with the COVID-19 vaccine to prevent one hospitalization across all subpopulations. NNV will be derived from the absolute risk reduction.

As a sensitivity analysis, we will estimate the share of COVID-19 hospitalizations that would be covered under alternative vaccination strategies, in which vaccination would be offered to all individuals with a NNV ≤ 500, 1000, 1500, or 2000. Coverage will be evaluated across VE values ranging from 30% to 65%, weighting by population size. Ninety-five percent confidence intervals will be derived using a multinomial bootstrap of subgroup-specific hospitalization counts, accounting for sampling variability while keeping subgroup membership fixed.

We will also calculate for different NNV cutoffs ranging from 500-2000, the proportion of the general population that would need to be vaccinated. CHS accounts for 52% of the population, but for a better representation of the total population, we will use data on age distribution from the Central Bureau of Statistics and adjust the required vaccination coverage to the general population^8^.

## Appendix B Additional results

**Figure S1.**
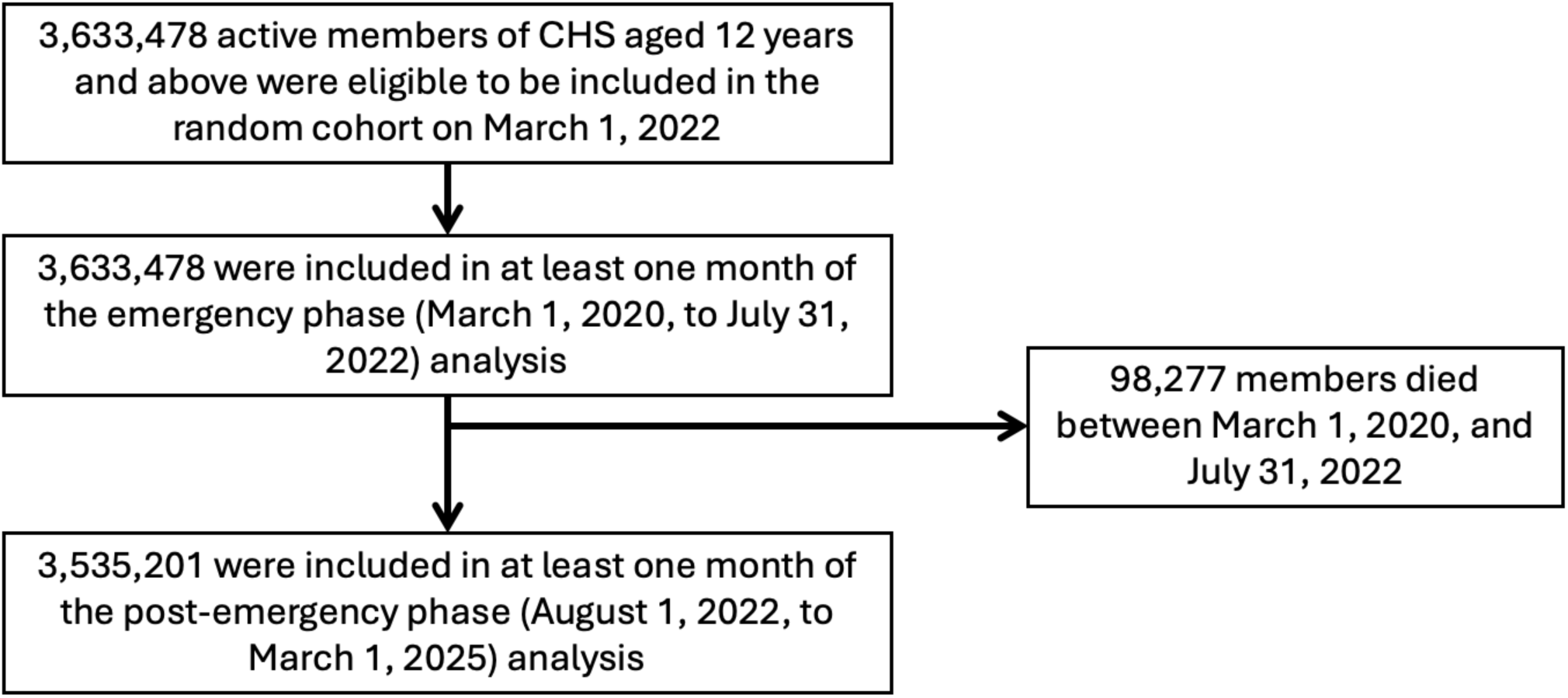
Study participants.

**Table S2.**
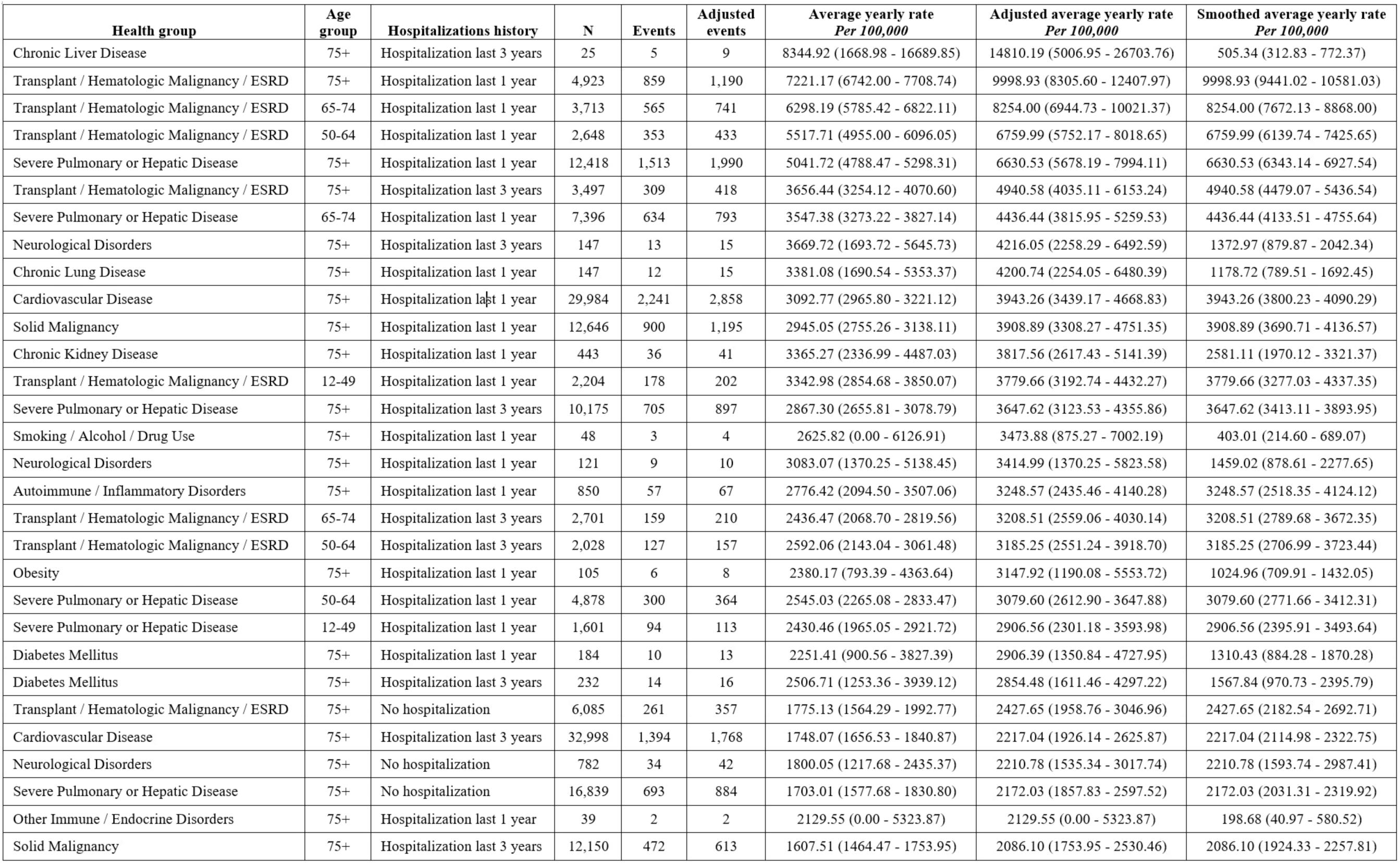
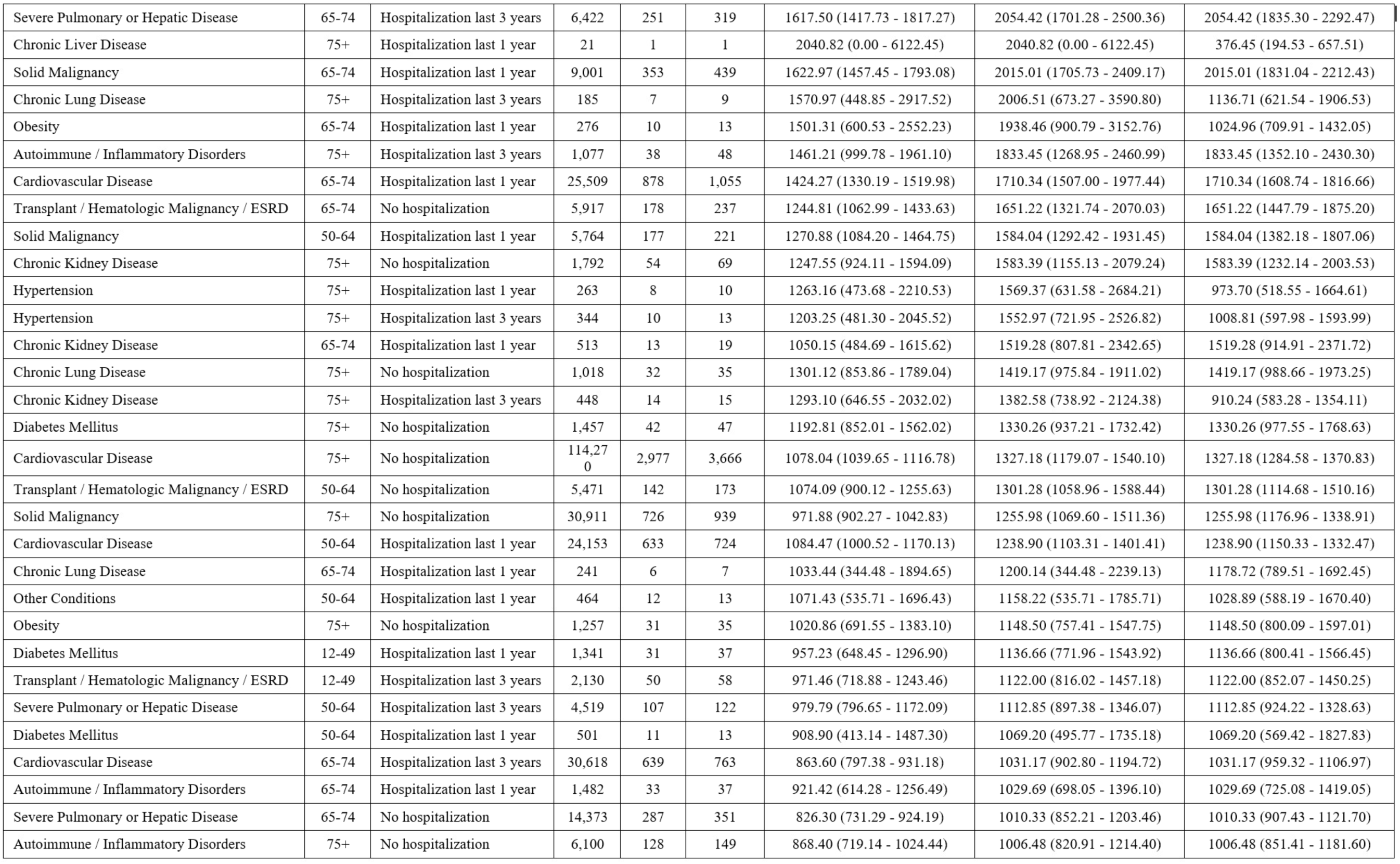
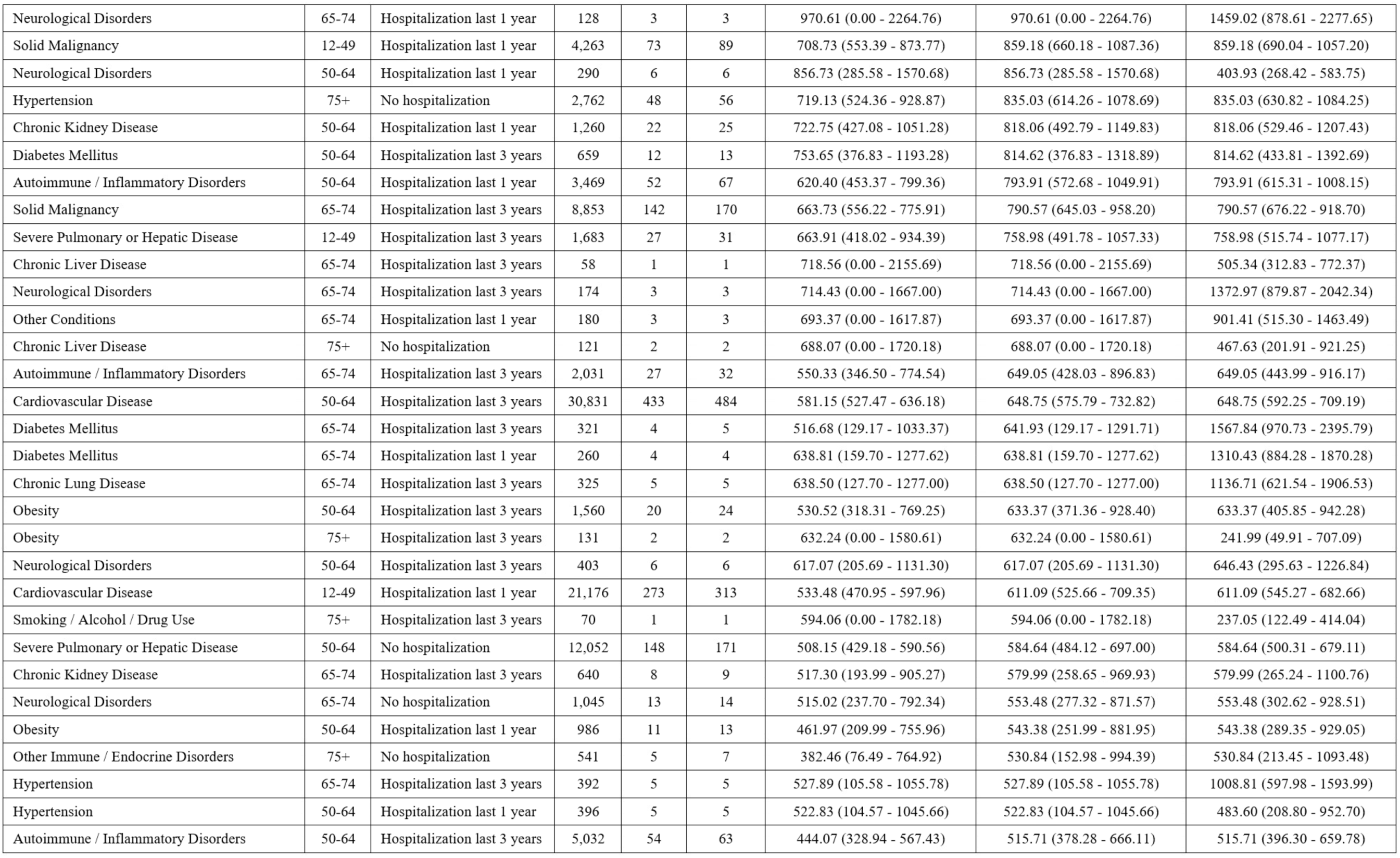
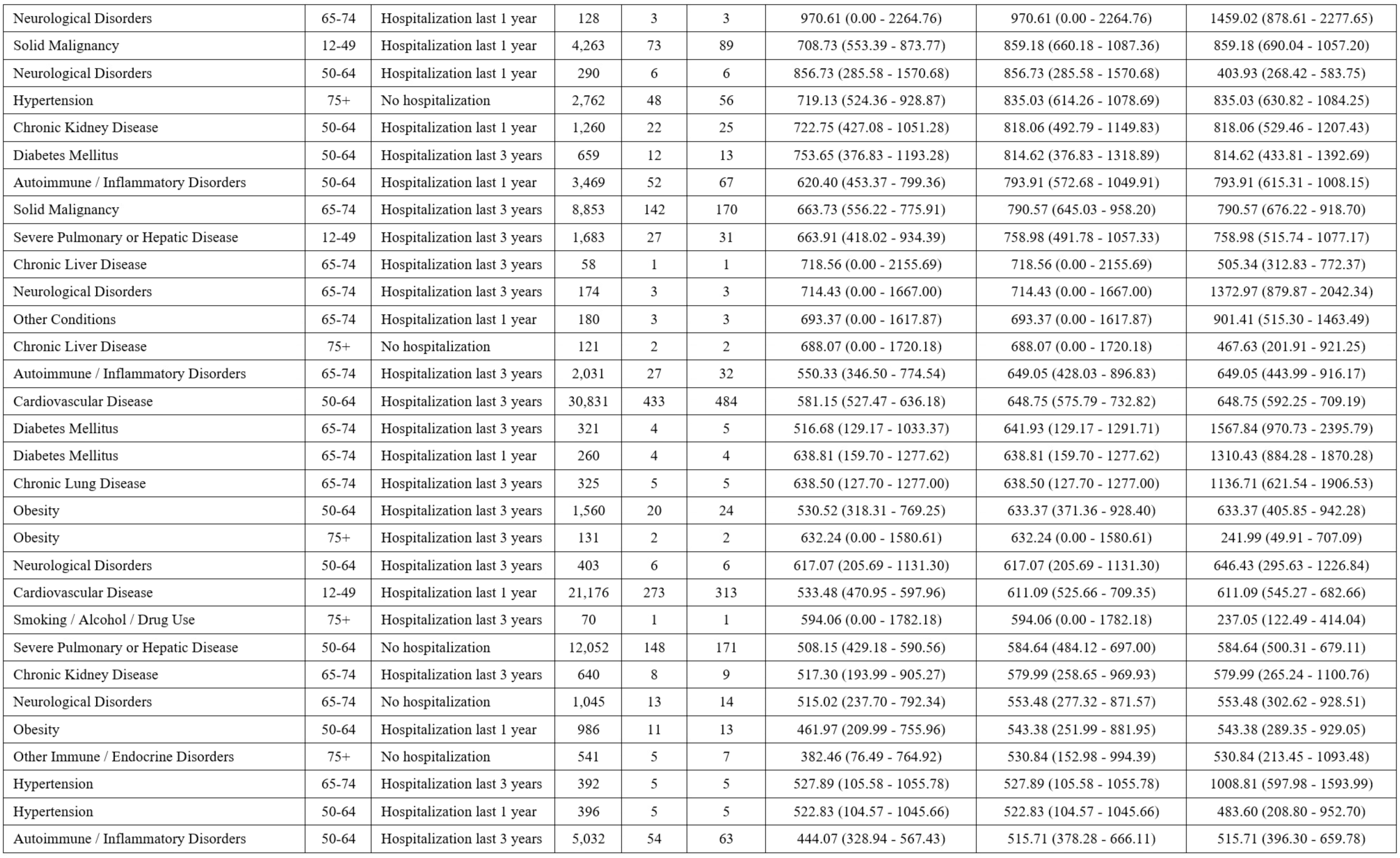
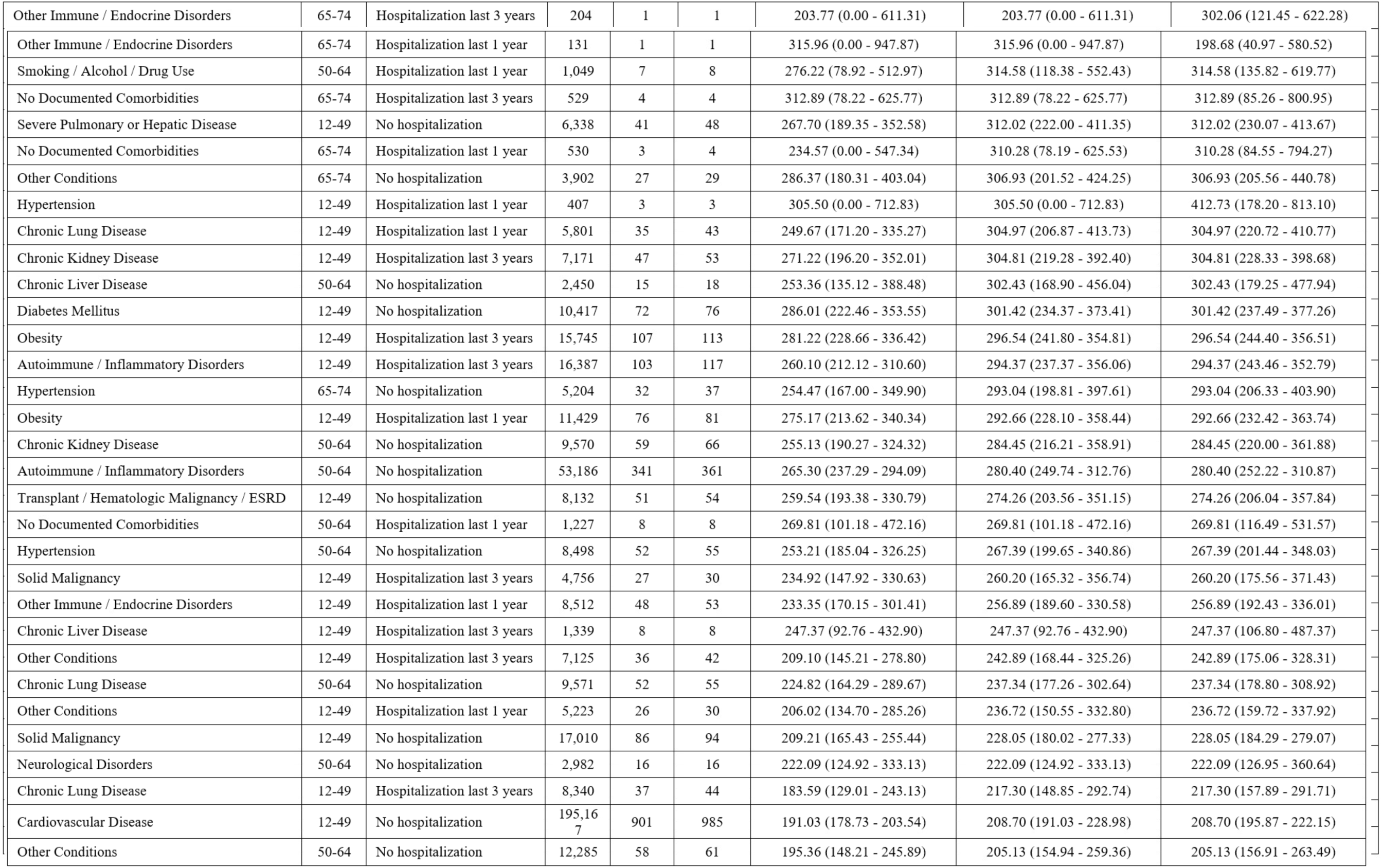
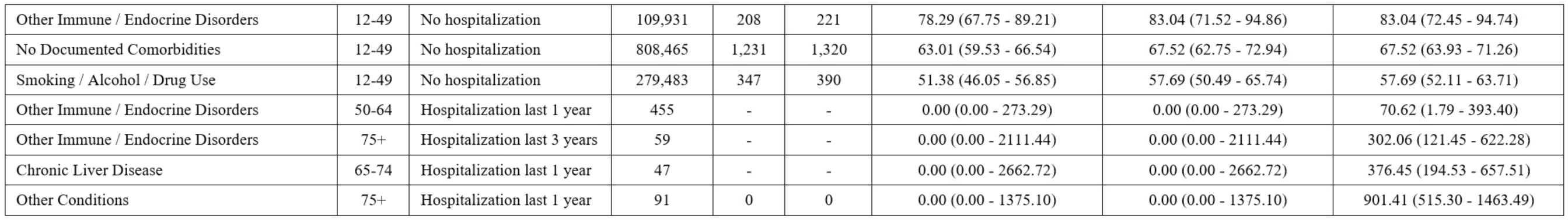
Average yearly rate per 100,000 individuals for each subgroup during the emergency phase.

**Table S3.** Average yearly rate per 100,000 individuals for each subgroup during the post-emergency phase.

| Health group | Age group | Hospitalizations history | N | Events | Adjusted events | Average yearly rate<br>Per 100,000 | Adjusted average yearly rate<br>Per 100,000 | Smoothed average yearly rate<br>Per 100,000 |
| --- | --- | --- | --- | --- | --- | --- | --- | --- |
| Transplant / Hematologic Malignancy / ESRD | 75+ | Hospitalization last 1 year | 5,694 | 471 | 491 | 3102.24 (2825.61 - 3385.46) | 3229.95 (2930.99 - 3536.95) | 3229.95 (2950.87 - 3528.25) |
| Transplant / Hematologic Malignancy / ESRD | 65-74 | Hospitalization last 1 year | 3,932 | 250 | 260 | 2384.68 (2098.52 - 2680.38) | 2476.82 (2174.83 - 2794.85) | 2476.82 (2185.14 - 2796.53) |
| Severe Pulmonary or Hepatic Disease | 75+ | Hospitalization last 1 year | 13,456 | 835 | 872 | 2327.17 (2171.10 - 2486.03) | 2429.89 (2251.92 - 2614.23) | 2429.89 (2271.41 - 2596.49) |
| Transplant / Hematologic Malignancy / ESRD | 50-64 | Hospitalization last 1 year | 2,780 | 111 | 112 | 1497.34 (1227.55 - 1780.62) | 1510.59 (1241.04 - 1794.11) | 1510.59 (1243.94 - 1817.40) |
| Smoking / Alcohol / Drug Use | 75+ | Hospitalization last 1 year | 52 | 2 | 2 | 1462.52 (0.00 - 3656.31) | 1462.52 (0.00 - 3656.31) | 185.42 (74.55 - 381.99) |
| Severe Pulmonary or Hepatic Disease | 65-74 | Hospitalization last 1 year | 8,066 | 282 | 288 | 1311.20 (1162.41 - 1464.63) | 1338.16 (1185.66 - 1497.18) | 1338.16 (1188.13 - 1501.87) |
| Neurological Disorders | 75+ | Hospitalization last 1 year | 141 | 5 | 5 | 1334.82 (266.96 - 2669.63) | 1334.82 (266.96 - 2669.63) | 609.17 (292.15 - 1120.04) |
| Chronic Kidney Disease | 75+ | Hospitalization last 1 year | 467 | 16 | 16 | 1285.31 (722.99 - 1927.97) | 1285.31 (722.99 - 1927.97) | 733.94 (448.36 - 1133.33) |
| Transplant / Hematologic Malignancy / ESRD | 75+ | Hospitalization last 3 years | 4,142 | 134 | 141 | 1213.30 (1014.10 - 1421.55) | 1274.75 (1059.37 - 1493.99) | 1274.75 (1073.12 - 1503.23) |
| Solid Malignancy | 75+ | Hospitalization last 1 year | 15,318 | 489 | 510 | 1197.20 (1091.92 - 1304.92) | 1247.07 (1133.54 - 1363.68) | 1247.07 (1141.22 - 1360.09) |
| Cardiovascular Disease | 75+ | Hospitalization last 1 year | 33,991 | 1,041 | 1,080 | 1148.49 (1078.98 - 1219.10) | 1191.25 (1114.29 - 1272.05) | 1191.25 (1121.29 - 1264.43) |
| Severe Pulmonary or Hepatic Disease | 75+ | Hospitalization last 3 years | 10,316 | 305 | 314 | 1108.79 (985.19 - 1236.03) | 1140.53 (1014.27 - 1272.38) | 1140.53 (1017.90 - 1273.85) |
| Other Immune / Endocrine Disorders | 75+ | Hospitalization last 3 years | 68 | 2 | 2 | 1110.08 (0.00 - 2775.21) | 1110.08 (0.00 - 2775.21) | 65.47 (7.93 - 236.48) |
| Severe Pulmonary or Hepatic Disease | 50-64 | Hospitalization last 1 year | 4,923 | 136 | 138 | 1036.07 (868.47 - 1211.28) | 1050.78 (876.08 - 1226.52) | 1050.78 (882.84 - 1241.34) |
| Hypertension | 75+ | Hospitalization last 1 year | 287 | 7 | 7 | 916.53 (261.87 - 1702.13) | 916.53 (261.87 - 1702.13) | 584.70 (267.39 - 1109.70) |
| Obesity | 75+ | Hospitalization last 1 year | 124 | 3 | 3 | 913.71 (0.00 - 2131.98) | 913.71 (0.00 - 2131.98) | 144.97 (53.20 - 315.53) |
| Transplant / Hematologic Malignancy / ESRD | 12-49 | Hospitalization last 1 year | 2,381 | 54 | 54 | 850.65 (630.11 - 1086.94) | 850.65 (630.11 - 1086.94) | 850.65 (639.09 - 1109.79) |
| Neurological Disorders | 75+ | No hospitalizations | 767 | 16 | 17 | 782.33 (440.06 - 1173.50) | 829.43 (440.06 - 1271.29) | 829.43 (483.24 - 1327.73) |
| Obesity | 75+ | Hospitalization last 3 years | 186 | 4 | 4 | 808.76 (202.19 - 1617.52) | 808.76 (202.19 - 1617.52) | 407.08 (163.68 - 838.58) |
| Severe Pulmonary or Hepatic Disease | 75+ | No hospitalizations | 15,955 | 324 | 337 | 761.52 (679.26 - 846.13) | 791.08 (705.11 - 881.39) | 791.08 (708.89 - 880.18) |
| Other Immune / Endocrine Disorders | 75+ | Hospitalization last 1 year | 51 | 1 | 1 | 747.66 (0.00 - 2242.99) | 747.66 (0.00 - 2242.99) | 159.50 (32.89 - 466.08) |
| Transplant / Hematologic Malignancy / ESRD | 65-74 | Hospitalization last 3 years | 2,853 | 54 | 55 | 709.85 (525.81 - 907.03) | 722.51 (538.96 - 920.17) | 722.51 (544.33 - 940.36) |
| Chronic Lung Disease | 75+ | Hospitalization last 3 years | 212 | 4 | 4 | 708.70 (177.17 - 1417.39) | 708.70 (177.17 - 1417.39) | 340.67 (110.62 - 794.88) |
| Transplant / Hematologic Malignancy / ESRD | 75+ | No hospitalizations | 6,252 | 111 | 116 | 665.80 (545.83 - 791.76) | 694.84 (569.83 - 827.75) | 694.84 (574.19 - 833.35) |
| Chronic Lung Disease | 75+ | Hospitalization last 1 year | 164 | 3 | 3 | 687.68 (0.00 - 1604.58) | 687.68 (0.00 - 1604.58) | 306.32 (140.08 - 581.43) |
| Autoimmune / Inflammatory Disorders | 75+ | Hospitalization last 1 year | 932 | 17 | 17 | 684.17 (362.21 - 1046.38) | 684.17 (362.21 - 1046.38) | 684.17 (398.60 - 1095.24) |
| Solid Malignancy | 75+ | Hospitalization last 3 years | 14,727 | 245 | 259 | 623.89 (547.50 - 702.84) | 658.46 (575.51 - 746.13) | 658.46 (580.72 - 743.70) |
| Cardiovascular Disease | 75+ | Hospitalization last 3 years | 36,866 | 610 | 634 | 620.50 (571.67 - 670.34) | 644.17 (591.00 - 698.82) | 644.17 (595.01 - 696.31) |
| Severe Pulmonary or Hepatic Disease | 12-49 | Hospitalization last 1 year | 1,695 | 26 | 28 | 575.48 (376.27 - 796.81) | 618.64 (398.41 - 863.21) | 618.64 (411.11 - 894.00) |
| Chronic Liver Disease | 65-74 | Hospitalization last 1 year | 62 | 1 | 1 | 609.45 (0.00 - 1828.34) | 609.45 (0.00 - 1828.34) | 227.29 (98.13 - 447.80) |
| Solid Malignancy | 65-74 | Hospitalization last 1 year | 9,399 | 145 | 150 | 578.55 (486.78 - 674.31) | 597.86 (502.74 - 698.25) | 597.86 (506.03 - 701.53) |
| Transplant / Hematologic Malignancy / ESRD | 50-64 | Hospitalization last 3 years | 2,182 | 29 | 30 | 498.53 (326.62 - 687.63) | 515.05 (343.81 - 704.82) | 515.05 (347.52 - 735.20) |
| Neurological Disorders | 75+ | Hospitalization last 3 years | 148 | 2 | 2 | 509.01 (0.00 - 1272.53) | 509.01 (0.00 - 1272.53) | 195.57 (53.29 - 500.69) |
| Severe Pulmonary or Hepatic Disease | 65-74 | Hospitalization last 3 years | 6,674 | 83 | 85 | 466.42 (370.89 - 567.57) | 477.34 (376.51 - 584.43) | 477.34 (381.30 - 590.21) |
| Smoking / Alcohol / Drug Use | 75+ | Hospitalization last 3 years | 82 | 1 | 1 | 458.02 (0.00 - 1374.05) | 458.02 (0.00 - 1374.05) | 32.44 (3.93 - 117.17) |
| Neurological Disorders | 50-64 | Hospitalization last 1 year | 330 | 4 | 4 | 455.54 (113.88 - 911.08) | 455.54 (113.88 - 911.08) | 108.12 (49.44 - 205.24) |
| Neurological Disorders | 65-74 | Hospitalization last 3 years | 170 | 2 | 2 | 442.15 (0.00 - 1105.38) | 442.15 (0.00 - 1105.38) | 195.57 (53.29 - 500.69) |
| Solid Malignancy | 75+ | No hospitalizations | 32,344 | 349 | 362 | 404.64 (362.90 - 447.54) | 419.25 (375.66 - 464.93) | 419.25 (377.18 - 464.73) |
| Diabetes Mellitus | 75+ | Hospitalization last 3 years | 183 | 2 | 2 | 409.84 (0.00 - 1024.59) | 409.84 (0.00 - 1024.59) | 252.41 (108.98 - 497.29) |
| Chronic Kidney Disease | 75+ | No hospitalizations | 1,769 | 17 | 19 | 360.53 (190.87 - 551.40) | 401.61 (233.29 - 593.82) | 401.61 (241.81 - 627.11) |
| Chronic Kidney Disease | 75+ | Hospitalization last 3 years | 466 | 4 | 5 | 322.49 (80.62 - 644.99) | 400.59 (80.62 - 806.23) | 214.43 (86.21 - 441.76) |
| Solid Malignancy | 50-64 | Hospitalization last 1 year | 6,028 | 62 | 62 | 385.71 (292.40 - 485.25) | 385.71 (292.40 - 485.25) | 385.71 (295.73 - 494.44) |
| Cardiovascular Disease | 65-74 | Hospitalization last 1 year | 26,522 | 268 | 272 | 378.94 (333.69 - 425.60) | 384.40 (339.35 - 431.26) | 384.40 (340.08 - 432.89) |
| Cardiovascular Disease | 75+ | No hospitalizations | 117,241 | 1,128 | 1,164 | 360.80 (340.01 - 381.91) | 372.25 (349.28 - 395.98) | 372.25 (351.17 - 394.26) |
| Diabetes Mellitus | 75+ | No hospitalizations | 1,212 | 12 | 12 | 371.40 (185.70 - 588.05) | 371.40 (185.70 - 588.05) | 371.40 (191.92 - 648.69) |
| Obesity | 65-74 | Hospitalization last 1 year | 315 | 3 | 3 | 357.28 (0.00 - 833.66) | 357.28 (0.00 - 833.66) | 144.97 (53.20 - 315.53) |
| Transplant / Hematologic Malignancy / ESRD | 65-74 | No hospitalizations | 5,560 | 50 | 52 | 337.26 (249.57 - 431.69) | 350.35 (256.32 - 451.93) | 350.35 (261.67 - 459.42) |
| Diabetes Mellitus | 65-74 | Hospitalization last 1 year | 221 | 2 | 2 | 340.04 (0.00 - 850.10) | 340.04 (0.00 - 850.10) | 272.10 (99.86 - 592.17) |
| Chronic Lung Disease | 50-64 | Hospitalization last 1 year | 688 | 6 | 6 | 327.42 (109.14 - 600.27) | 327.42 (109.14 - 600.27) | 327.42 (120.17 - 712.54) |
| Severe Pulmonary or Hepatic Disease | 50-64 | Hospitalization last 3 years | 4,356 | 35 | 36 | 301.33 (206.63 - 404.65) | 309.60 (215.24 - 413.26) | 309.60 (216.85 - 428.60) |
| Smoking / Alcohol / Drug Use | 65-74 | Hospitalization last 1 year | 245 | 2 | 2 | 306.47 (0.00 - 766.19) | 306.47 (0.00 - 766.19) | 185.42 (74.55 - 381.99) |
| Chronic Lung Disease | 75+ | No hospitalizations | 1,004 | 7 | 8 | 261.70 (74.77 - 486.01) | 298.01 (112.16 - 523.40) | 298.01 (128.67 - 587.12) |
| Severe Pulmonary or Hepatic Disease | 65-74 | No hospitalizations | 13,639 | 101 | 102 | 277.71 (225.47 - 332.70) | 280.37 (228.22 - 335.45) | 280.37 (228.61 - 340.34) |
| Chronic Liver Disease | 75+ | No hospitalizations | 136 | 1 | 1 | 275.80 (0.00 - 827.40) | 275.80 (0.00 - 827.40) | 53.42 (1.35 - 297.61) |
| Chronic Kidney Disease | 65-74 | Hospitalization last 1 year | 556 | 4 | 4 | 270.24 (67.56 - 540.48) | 270.24 (67.56 - 540.48) | 270.24 (73.64 - 691.80) |
| Hypertension | 65-74 | Hospitalization last 1 year | 291 | 2 | 2 | 257.90 (0.00 - 644.75) | 257.90 (0.00 - 644.75) | 584.70 (267.39 - 1109.70) |
| Neurological Disorders | 65-74 | Hospitalization last 1 year | 146 | 1 | 1 | 257.12 (0.00 - 771.37) | 257.12 (0.00 - 771.37) | 609.17 (292.15 - 1120.04) |
| Diabetes Mellitus | 75+ | Hospitalization last 1 year | 150 | 1 | 1 | 251.57 (0.00 - 754.72) | 251.57 (0.00 - 754.72) | 272.10 (99.86 - 592.17) |
| Diabetes Mellitus | 50-64 | Hospitalization last 1 year | 458 | 3 | 3 | 246.02 (0.00 - 574.04) | 246.02 (0.00 - 574.04) | 276.61 (89.82 - 645.42) |
| Obesity | 65-74 | Hospitalization last 3 years | 460 | 3 | 3 | 244.90 (0.00 - 571.43) | 244.90 (0.00 - 571.43) | 407.08 (163.68 - 838.58) |
| Diabetes Mellitus | 65-74 | Hospitalization last 3 years | 307 | 2 | 2 | 244.45 (0.00 - 611.12) | 244.45 (0.00 - 611.12) | 252.41 (108.98 - 497.29) |
| Hypertension | 75+ | No hospitalizations | 2,598 | 16 | 16 | 230.95 (129.91 - 346.43) | 230.95 (129.91 - 346.43) | 230.95 (132.01 - 375.03) |
| Chronic Liver Disease | 12-49 | Hospitalization last 1 year | 1,007 | 6 | 6 | 223.52 (74.51 - 409.79) | 223.52 (74.51 - 409.79) | 223.52 (82.03 - 486.45) |
| Other Conditions | 65-74 | Hospitalization last 1 year | 172 | 1 | 1 | 218.66 (0.00 - 655.98) | 218.66 (0.00 - 655.98) | 51.06 (1.29 - 284.43) |
| Diabetes Mellitus | 50-64 | Hospitalization last 3 years | 699 | 4 | 4 | 214.67 (53.67 - 429.34) | 214.67 (53.67 - 429.34) | 214.67 (58.49 - 549.56) |
| Cardiovascular Disease | 50-64 | Hospitalization last 1 year | 24,518 | 133 | 135 | 203.43 (169.78 - 238.61) | 206.41 (172.84 - 241.67) | 206.41 (173.06 - 244.31) |
| Chronic Kidney Disease | 50-64 | Hospitalization last 1 year | 1,410 | 7 | 7 | 186.26 (53.22 - 345.91) | 186.26 (53.22 - 345.91) | 186.26 (74.89 - 383.74) |
| Hypertension | 65-74 | Hospitalization last 3 years | 411 | 2 | 2 | 182.70 (0.00 - 456.76) | 182.70 (0.00 - 456.76) | 146.89 (30.29 - 429.24) |
| Neurological Disorders | 65-74 | No hospitalizations | 1,039 | 5 | 5 | 180.61 (36.12 - 361.23) | 180.61 (36.12 - 361.23) | 180.61 (58.65 - 421.44) |
| Severe Pulmonary or Hepatic Disease | 12-49 | Hospitalization last 3 years | 1,661 | 7 | 8 | 158.11 (45.17 - 293.63) | 179.91 (67.76 - 316.22) | 179.91 (77.67 - 354.47) |
| Autoimmune / Inflammatory Disorders | 75+ | Hospitalization last 3 years | 1,263 | 5 | 6 | 148.46 (29.69 - 296.93) | 177.29 (59.39 - 326.62) | 177.29 (65.07 - 385.86) |
| Cardiovascular Disease | 65-74 | Hospitalization last 3 years | 33,196 | 151 | 154 | 170.58 (143.47 - 198.82) | 173.88 (146.86 - 202.21) | 173.88 (147.50 - 203.61) |
| Solid Malignancy | 12-49 | Hospitalization last 1 year | 4,582 | 21 | 21 | 171.90 (106.42 - 245.58) | 171.90 (106.42 - 245.58) | 171.90 (106.41 - 262.76) |
| Autoimmune / Inflammatory Disorders | 65-74 | Hospitalization last 3 years | 2,214 | 10 | 10 | 169.43 (67.77 - 288.03) | 169.43 (67.77 - 288.03) | 169.43 (81.25 - 311.57) |
| Autoimmune / Inflammatory Disorders | 75+ | No hospitalizations | 6,497 | 28 | 29 | 161.63 (103.91 - 225.13) | 167.28 (109.68 - 230.90) | 167.28 (112.03 - 240.24) |
| Autoimmune / Inflammatory Disorders | 65-74 | Hospitalization last 1 year | 1,576 | 7 | 7 | 166.64 (47.61 - 309.48) | 166.64 (47.61 - 309.48) | 166.64 (67.00 - 343.32) |
| Chronic Liver Disease | 50-64 | Hospitalization last 1 year | 233 | 1 | 1 | 161.10 (0.00 - 483.29) | 161.10 (0.00 - 483.29) | 230.60 (99.56 - 454.34) |
| Solid Malignancy | 65-74 | Hospitalization last 3 years | 9,700 | 39 | 41 | 150.79 (104.39 - 201.05) | 158.23 (112.12 - 208.78) | 158.23 (113.55 - 214.65) |
| Other Immune / Endocrine Disorders | 50-64 | Hospitalization last 1 year | 501 | 2 | 2 | 149.95 (0.00 - 374.88) | 149.95 (0.00 - 374.88) | 149.95 (18.16 - 541.59) |
| Severe Pulmonary or Hepatic Disease | 50-64 | No hospitalizations | 10,823 | 36 | 37 | 124.74 (86.63 - 166.33) | 128.11 (90.09 - 169.79) | 128.11 (90.20 - 176.58) |
| Transplant / Hematologic Malignancy / ESRD | 12-49 | Hospitalization last 3 years | 2,265 | 7 | 7 | 115.91 (33.12 - 215.26) | 115.91 (33.12 - 215.26) | 115.91 (46.60 - 238.81) |
| Solid Malignancy | 50-64 | Hospitalization last 3 years | 6,616 | 20 | 20 | 113.38 (68.03 - 164.40) | 113.38 (68.03 - 164.40) | 113.38 (69.26 - 175.10) |
| Chronic Lung Disease | 65-74 | Hospitalization last 3 years | 339 | 1 | 1 | 110.71 (0.00 - 332.13) | 110.71 (0.00 - 332.13) | 340.67 (110.62 - 794.88) |
| Chronic Kidney Disease | 50-64 | Hospitalization last 3 years | 2,066 | 6 | 6 | 108.92 (36.31 - 199.68) | 108.92 (36.31 - 199.68) | 108.92 (39.97 - 237.06) |
| Chronic Kidney Disease | 12-49 | Hospitalization last 1 year | 5,940 | 17 | 17 | 107.34 (56.83 - 164.16) | 107.34 (56.83 - 164.16) | 107.34 (62.53 - 171.86) |
| Hypertension | 75+ | Hospitalization last 3 years | 356 | 1 | 1 | 105.53 (0.00 - 316.59) | 105.53 (0.00 - 316.59) | 146.89 (30.29 - 429.24) |
| Smoking / Alcohol / Drug Use | 50-64 | Hospitalization last 1 year | 1,120 | 3 | 3 | 100.47 (0.00 - 234.42) | 100.47 (0.00 - 234.42) | 100.47 (20.72 - 293.59) |
| Solid Malignancy | 65-74 | No hospitalizations | 27,438 | 70 | 73 | 95.67 (73.80 - 118.91) | 99.63 (77.90 - 123.01) | 99.63 (78.09 - 125.27) |
| Chronic Kidney Disease | 65-74 | Hospitalization last 3 years | 757 | 2 | 2 | 99.19 (0.00 - 247.97) | 99.19 (0.00 - 247.97) | 99.19 (12.01 - 358.27) |
| Cardiovascular Disease | 12-49 | Hospitalization last 1 year | 22,240 | 56 | 56 | 94.42 (70.82 - 119.72) | 94.42 (70.82 - 119.72) | 94.42 (71.32 - 122.61) |
| Hypertension | 12-49 | Hospitalization last 1 year | 406 | 1 | 1 | 92.57 (0.00 - 277.71) | 92.57 (0.00 - 277.71) | 46.93 (1.19 - 261.44) |
| Diabetes Mellitus | 12-49 | Hospitalization last 3 years | 1,806 | 4 | 4 | 83.06 (20.77 - 166.12) | 83.06 (20.77 - 166.12) | 83.06 (22.63 - 212.65) |
| Autoimmune / Inflammatory Disorders | 12-49 | Hospitalization last 1 year | 11,487 | 25 | 25 | 81.62 (52.23 - 114.26) | 81.62 (52.23 - 114.26) | 81.62 (52.82 - 120.49) |
| Chronic Lung Disease | 12-49 | Hospitalization last 1 year | 6,031 | 13 | 13 | 80.84 (37.31 - 130.58) | 80.84 (37.31 - 130.58) | 80.84 (43.04 - 138.24) |
| Chronic Kidney Disease | 65-74 | No hospitalizations | 3,729 | 8 | 8 | 80.46 (30.17 - 140.81) | 80.46 (30.17 - 140.81) | 80.46 (34.74 - 158.53) |
| Obesity | 75+ | No hospitalizations | 1,417 | 3 | 3 | 79.41 (0.00 - 185.28) | 79.41 (0.00 - 185.28) | 79.41 (16.38 - 232.06) |
| Cardiovascular Disease | 50-64 | Hospitalization last 3 years | 32,437 | 61 | 63 | 70.52 (53.18 - 89.02) | 72.75 (55.49 - 91.33) | 72.75 (55.90 - 93.08) |
| Autoimmune / Inflammatory Disorders | 50-64 | Hospitalization last 1 year | 3,678 | 7 | 7 | 71.38 (20.39 - 132.57) | 71.38 (20.39 - 132.57) | 71.38 (28.70 - 147.07) |
| Cardiovascular Disease | 65-74 | No hospitalizations | 159,569 | 279 | 288 | 65.57 (58.05 - 73.32) | 67.62 (59.69 - 75.67) | 67.62 (60.04 - 75.90) |
| Transplant / Hematologic Malignancy / ESRD | 50-64 | No hospitalizations | 5,577 | 10 | 10 | 67.24 (26.90 - 114.32) | 67.24 (26.90 - 114.32) | 67.24 (32.24 - 123.65) |
| Severe Pulmonary or Hepatic Disease | 12-49 | No hospitalizations | 6,134 | 10 | 10 | 61.14 (24.46 - 103.94) | 61.14 (24.46 - 103.94) | 61.14 (29.32 - 112.44) |
| No Documented Comorbidities | 65-74 | Hospitalization last 3 years | 618 | 1 | 1 | 60.72 (0.00 - 182.16) | 60.72 (0.00 - 182.16) | 60.72 (1.54 - 338.27) |
| No Documented Comorbidities | 65-74 | Hospitalization last 1 year | 624 | 1 | 1 | 60.11 (0.00 - 180.33) | 60.11 (0.00 - 180.33) | 60.11 (1.52 - 334.88) |
| Other Immune / Endocrine Disorders | 75+ | No hospitalizations | 643 | 1 | 1 | 58.32 (0.00 - 174.97) | 58.32 (0.00 - 174.97) | 58.32 (1.48 - 324.90) |
| Chronic Lung Disease | 65-74 | No hospitalizations | 2,635 | 4 | 4 | 56.95 (14.24 - 113.89) | 56.95 (14.24 - 113.89) | 56.95 (15.52 - 145.81) |
| Other Conditions | 75+ | No hospitalizations | 1,319 | 2 | 2 | 56.87 (0.00 - 142.16) | 56.87 (0.00 - 142.16) | 56.87 (6.89 - 205.42) |
| Neurological Disorders | 12-49 | Hospitalization last 1 year | 2,647 | 4 | 4 | 56.68 (14.17 - 113.37) | 56.68 (14.17 - 113.37) | 56.68 (15.44 - 145.12) |
| Diabetes Mellitus | 12-49 | Hospitalization last 1 year | 1,359 | 2 | 2 | 55.20 (0.00 - 138.00) | 55.20 (0.00 - 138.00) | 55.20 (6.69 - 199.39) |
| Autoimmune / Inflammatory Disorders | 50-64 | Hospitalization last 3 years | 5,648 | 8 | 8 | 53.12 (19.92 - 92.96) | 53.12 (19.92 - 92.96) | 53.12 (22.93 - 104.67) |
| Chronic Liver Disease | 12-49 | Hospitalization last 3 years | 1,469 | 2 | 2 | 51.07 (0.00 - 127.67) | 51.07 (0.00 - 127.67) | 51.07 (6.18 - 184.47) |
| Diabetes Mellitus | 65-74 | No hospitalizations | 2,997 | 4 | 4 | 50.05 (12.51 - 100.10) | 50.05 (12.51 - 100.10) | 50.05 (13.64 - 128.14) |
| Other Conditions | 12-49 | Hospitalization last 1 year | 5,364 | 7 | 7 | 48.94 (13.98 - 90.89) | 48.94 (13.98 - 90.89) | 48.94 (19.68 - 100.83) |
| Neurological Disorders | 50-64 | No hospitalizations | 3,175 | 4 | 4 | 47.25 (11.81 - 94.50) | 47.25 (11.81 - 94.50) | 47.25 (12.87 - 120.98) |
| Obesity | 65-74 | No hospitalizations | 5,830 | 7 | 7 | 45.03 (12.87 - 83.63) | 45.03 (12.87 - 83.63) | 45.03 (18.10 - 92.78) |
| Cardiovascular Disease | 12-49 | Hospitalization last 3 years | 30,954 | 34 | 34 | 41.19 (27.86 - 55.73) | 41.19 (27.86 - 55.73) | 41.19 (28.53 - 57.56) |
| Solid Malignancy | 12-49 | Hospitalization last 3 years | 5,577 | 6 | 6 | 40.35 (13.45 - 73.97) | 40.35 (13.45 - 73.97) | 40.35 (14.81 - 87.82) |
| Autoimmune / Inflammatory Disorders | 65-74 | No hospitalizations | 17,877 | 19 | 19 | 39.86 (23.07 - 58.74) | 39.86 (23.07 - 58.74) | 39.86 (24.00 - 62.25) |
| Other Conditions | 65-74 | No hospitalizations | 3,829 | 4 | 4 | 39.18 (9.80 - 78.37) | 39.18 (9.80 - 78.37) | 39.18 (10.68 - 100.31) |
| Smoking / Alcohol / Drug Use | 75+ | No hospitalizations | 978 | 1 | 1 | 38.38 (0.00 - 115.14) | 38.38 (0.00 - 115.14) | 38.38 (0.97 - 213.83) |
| Chronic Lung Disease | 50-64 | Hospitalization last 3 years | 1,051 | 1 | 1 | 35.70 (0.00 - 107.11) | 35.70 (0.00 - 107.11) | 35.70 (0.90 - 198.90) |
| Solid Malignancy | 50-64 | No hospitalizations | 22,103 | 20 | 20 | 33.93 (20.36 - 49.20) | 33.93 (20.36 - 49.20) | 33.93 (20.73 - 52.40) |
| Other Immune / Endocrine Disorders | 12-49 | Hospitalization last 3 years | 13,631 | 12 | 12 | 33.01 (16.51 - 52.27) | 33.01 (16.51 - 52.27) | 33.01 (17.06 - 57.66) |
| Chronic Lung Disease | 50-64 | No hospitalizations | 10,318 | 9 | 9 | 32.71 (14.54 - 54.52) | 32.71 (14.54 - 54.52) | 32.71 (14.96 - 62.09) |
| Autoimmune / Inflammatory Disorders | 12-49 | Hospitalization last 3 years | 17,382 | 15 | 15 | 32.36 (17.26 - 49.62) | 32.36 (17.26 - 49.62) | 32.36 (18.11 - 53.37) |
| Chronic Kidney Disease | 12-49 | Hospitalization last 3 years | 8,116 | 7 | 7 | 32.35 (9.24 - 60.07) | 32.35 (9.24 - 60.07) | 32.35 (13.01 - 66.65) |
| Transplant / Hematologic Malignancy / ESRD | 12-49 | No hospitalizations | 8,116 | 7 | 7 | 32.34 (9.24 - 60.07) | 32.34 (9.24 - 60.07) | 32.34 (13.00 - 66.63) |
| Smoking / Alcohol / Drug Use | 12-49 | Hospitalization last 1 year | 12,816 | 11 | 11 | 32.19 (14.63 - 52.67) | 32.19 (14.63 - 52.67) | 32.19 (16.07 - 57.60) |
| Other Immune / Endocrine Disorders | 12-49 | Hospitalization last 1 year | 9,334 | 8 | 8 | 32.14 (12.05 - 56.25) | 32.14 (12.05 - 56.25) | 32.14 (13.88 - 63.33) |
| Neurological Disorders | 12-49 | Hospitalization last 3 years | 3,544 | 2 | 3 | 21.17 (0.00 - 52.91) | 31.43 (0.00 - 74.08) | 31.43 (6.48 - 91.85) |
| Obesity | 12-49 | Hospitalization last 1 year | 12,989 | 10 | 10 | 28.87 (11.55 - 49.08) | 28.87 (11.55 - 49.08) | 28.87 (13.84 - 53.09) |
| Cardiovascular Disease | 50-64 | No hospitalizations | 204,184 | 138 | 141 | 25.34 (21.12 - 29.57) | 25.88 (21.67 - 30.30) | 25.88 (21.78 - 30.52) |
| Chronic Lung Disease | 12-49 | Hospitalization last 3 years | 8,847 | 6 | 6 | 25.43 (8.48 - 46.63) | 25.43 (8.48 - 46.63) | 25.43 (9.33 - 55.35) |
| No Documented Comorbidities | 75+ | No hospitalizations | 7,478 | 5 | 5 | 25.08 (5.02 - 50.15) | 25.08 (5.02 - 50.15) | 25.08 (8.14 - 58.53) |
| Hypertension | 65-74 | No hospitalizations | 4,655 | 3 | 3 | 24.17 (0.00 - 56.39) | 24.17 (0.00 - 56.39) | 24.17 (4.98 - 70.63) |
| No Documented Comorbidities | 12-49 | Hospitalization last 3 years | 40,241 | 23 | 23 | 21.43 (13.05 - 30.75) | 21.43 (13.05 - 30.75) | 21.43 (13.58 - 32.16) |
| Smoking / Alcohol / Drug Use | 50-64 | Hospitalization last 3 years | 1,830 | 1 | 1 | 20.50 (0.00 - 61.49) | 20.50 (0.00 - 61.49) | 20.50 (0.52 - 114.21) |
| Diabetes Mellitus | 50-64 | No hospitalizations | 7,346 | 4 | 4 | 20.42 (5.11 - 40.84) | 20.42 (5.11 - 40.84) | 20.42 (5.56 - 52.28) |
| Neurological Disorders | 12-49 | No hospitalizations | 18,504 | 10 | 10 | 20.27 (8.11 - 34.45) | 20.27 (8.11 - 34.45) | 20.27 (9.72 - 37.28) |
| Obesity | 12-49 | Hospitalization last 3 years | 18,962 | 10 | 10 | 19.78 (7.91 - 33.62) | 19.78 (7.91 - 33.62) | 19.78 (9.49 - 36.38) |
| No Documented Comorbidities | 12-49 | Hospitalization last 1 year | 27,264 | 14 | 14 | 19.26 (9.63 - 30.26) | 19.26 (9.63 - 30.26) | 19.26 (10.53 - 32.31) |
| Solid Malignancy | 12-49 | No hospitalizations | 17,813 | 9 | 9 | 18.95 (8.42 - 31.58) | 18.95 (8.42 - 31.58) | 18.95 (8.67 - 35.97) |
| Smoking / Alcohol / Drug Use | 65-74 | No hospitalizations | 6,129 | 3 | 3 | 18.36 (0.00 - 42.83) | 18.36 (0.00 - 42.83) | 18.36 (3.79 - 53.66) |
| Chronic Kidney Disease | 12-49 | No hospitalizations | 36,574 | 14 | 15 | 14.35 (7.18 - 22.56) | 15.34 (8.20 - 23.58) | 15.34 (8.59 - 25.30) |
| Chronic Liver Disease | 50-64 | No hospitalizations | 2,578 | 1 | 1 | 14.55 (0.00 - 43.64) | 14.55 (0.00 - 43.64) | 14.55 (0.37 - 81.07) |
| Other Immune / Endocrine Disorders | 65-74 | No hospitalizations | 2,627 | 1 | 1 | 14.28 (0.00 - 42.84) | 14.28 (0.00 - 42.84) | 14.28 (0.36 - 79.56) |
| Hypertension | 50-64 | No hospitalizations | 8,170 | 3 | 3 | 13.77 (0.00 - 32.13) | 13.77 (0.00 - 32.13) | 13.77 (2.84 - 40.24) |
| Autoimmune / Inflammatory Disorders | 50-64 | No hospitalizations | 55,185 | 20 | 20 | 13.59 (8.15 - 19.71) | 13.59 (8.15 - 19.71) | 13.59 (8.30 - 20.99) |
| Cardiovascular Disease | 12-49 | No hospitalizations | 192,909 | 62 | 62 | 12.05 (9.14 - 15.16) | 12.05 (9.14 - 15.16) | 12.05 (9.24 - 15.45) |
| Autoimmune / Inflammatory Disorders | 12-49 | No hospitalizations | 131,080 | 41 | 41 | 11.73 (8.30 - 15.45) | 11.73 (8.30 - 15.45) | 11.73 (8.42 - 15.91) |
| Chronic Lung Disease | 12-49 | No hospitalizations | 89,007 | 27 | 27 | 11.38 (7.16 - 16.01) | 11.38 (7.16 - 16.01) | 11.38 (7.50 - 16.56) |
| Diabetes Mellitus | 12-49 | No hospitalizations | 10,529 | 3 | 3 | 10.69 (0.00 - 24.93) | 10.69 (0.00 - 24.93) | 10.69 (2.20 - 31.24) |
| No Documented Comorbidities | 65-74 | No hospitalizations | 12,109 | 3 | 3 | 9.29 (0.00 - 21.68) | 9.29 (0.00 - 21.68) | 9.29 (1.92 - 27.15) |
| Other Immune / Endocrine Disorders | 12-49 | No hospitalizations | 114,276 | 28 | 28 | 9.19 (5.91 - 12.80) | 9.19 (5.91 - 12.80) | 9.19 (6.11 - 13.28) |
| Other Immune / Endocrine Disorders | 50-64 | No hospitalizations | 12,265 | 3 | 3 | 9.17 (0.00 - 21.40) | 9.17 (0.00 - 21.40) | 9.17 (1.89 - 26.80) |
| No Documented Comorbidities | 75+ | Hospitalization last 1 year | 1,259 | 0 | 0 | 0.00 (0.00 - 89.39) | 0.00 (0.00 - 89.39) | 0.00 (0.00 - 109.87) |
| Obesity | 50-64 | Hospitalization last 3 years | 1,842 | 0 | 0 | 0.00 (0.00 - 61.10) | 0.00 (0.00 - 61.10) | 0.00 (0.00 - 75.10) |
| No Documented Comorbidities | 50-64 | Hospitalization last 1 year | 1,347 | 0 | 0 | 0.00 (0.00 - 83.57) | 0.00 (0.00 - 83.57) | 0.00 (0.00 - 102.69) |
| Obesity | 50-64 | Hospitalization last 1 year | 1,114 | 0 | 0 | 0.00 (0.00 - 101.07) | 0.00 (0.00 - 101.07) | 0.00 (0.00 - 124.17) |
| Chronic Liver Disease | 65-74 | Hospitalization last 3 years | 90 | 0 | 0 | 0.00 (0.00 - 1257.86) | 0.00 (0.00 - 1257.86) | 38.83 (4.70 - 140.26) |
| Other Conditions | 75+ | Hospitalization last 3 years | 142 | 0 | 0 | 0.00 (0.00 - 796.11) | 0.00 (0.00 - 796.11) | 0.00 (0.00 - 112.10) |
| Other Immune / Endocrine Disorders | 65-74 | Hospitalization last 1 year | 155 | 0 | 0 | 0.00 (0.00 - 726.25) | 0.00 (0.00 - 726.25) | 159.50 (32.89 - 466.08) |

**Table S4.**
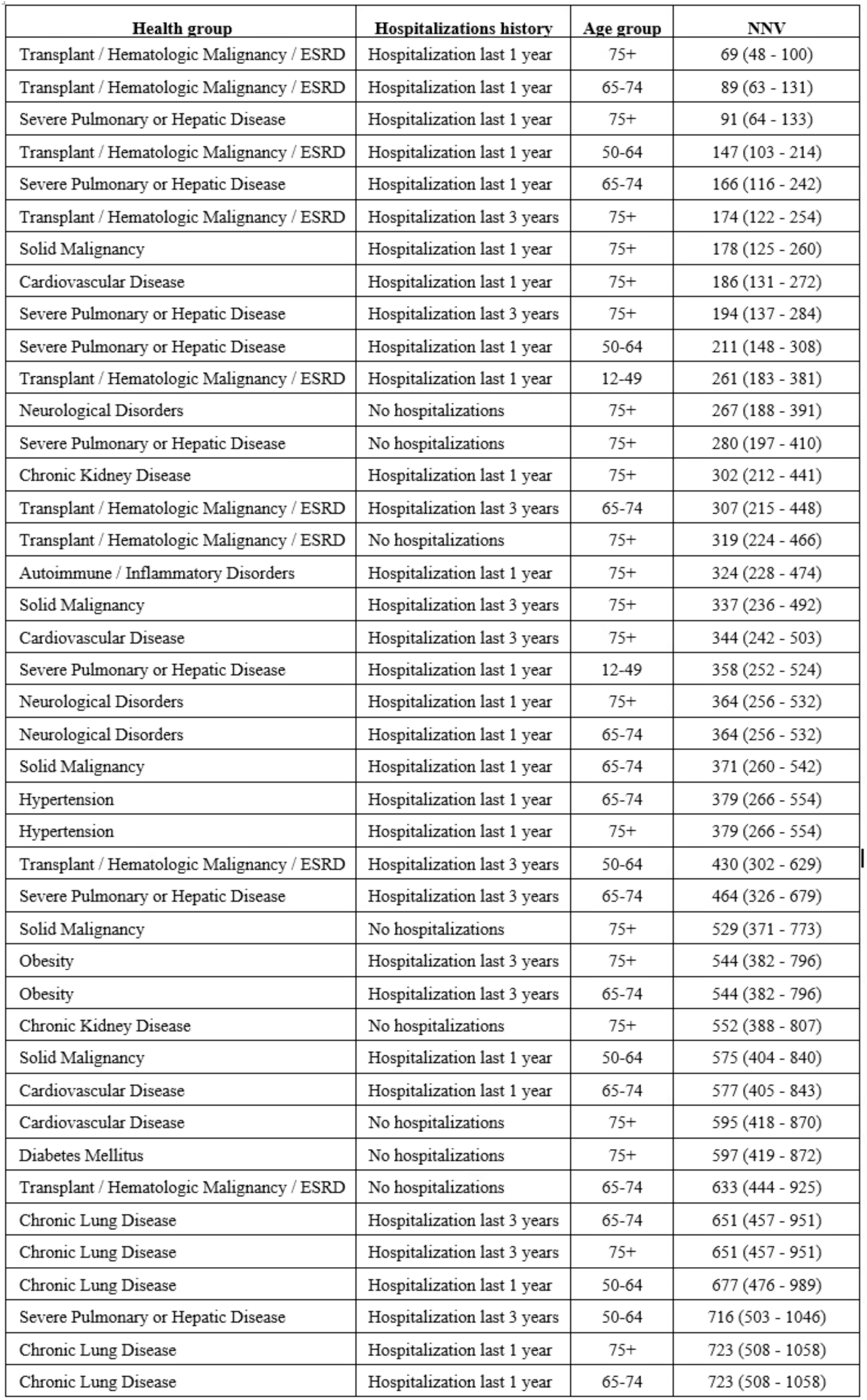
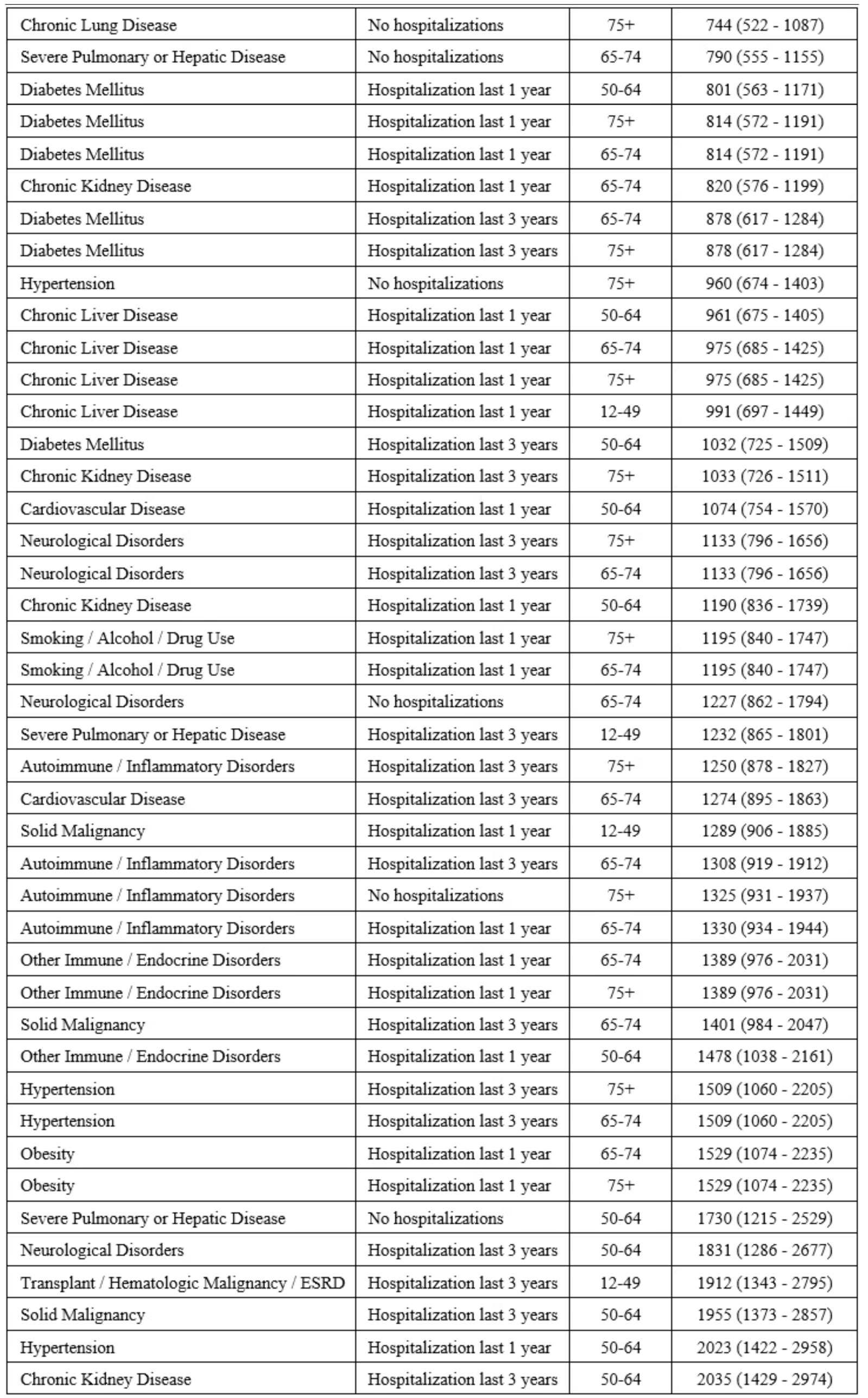
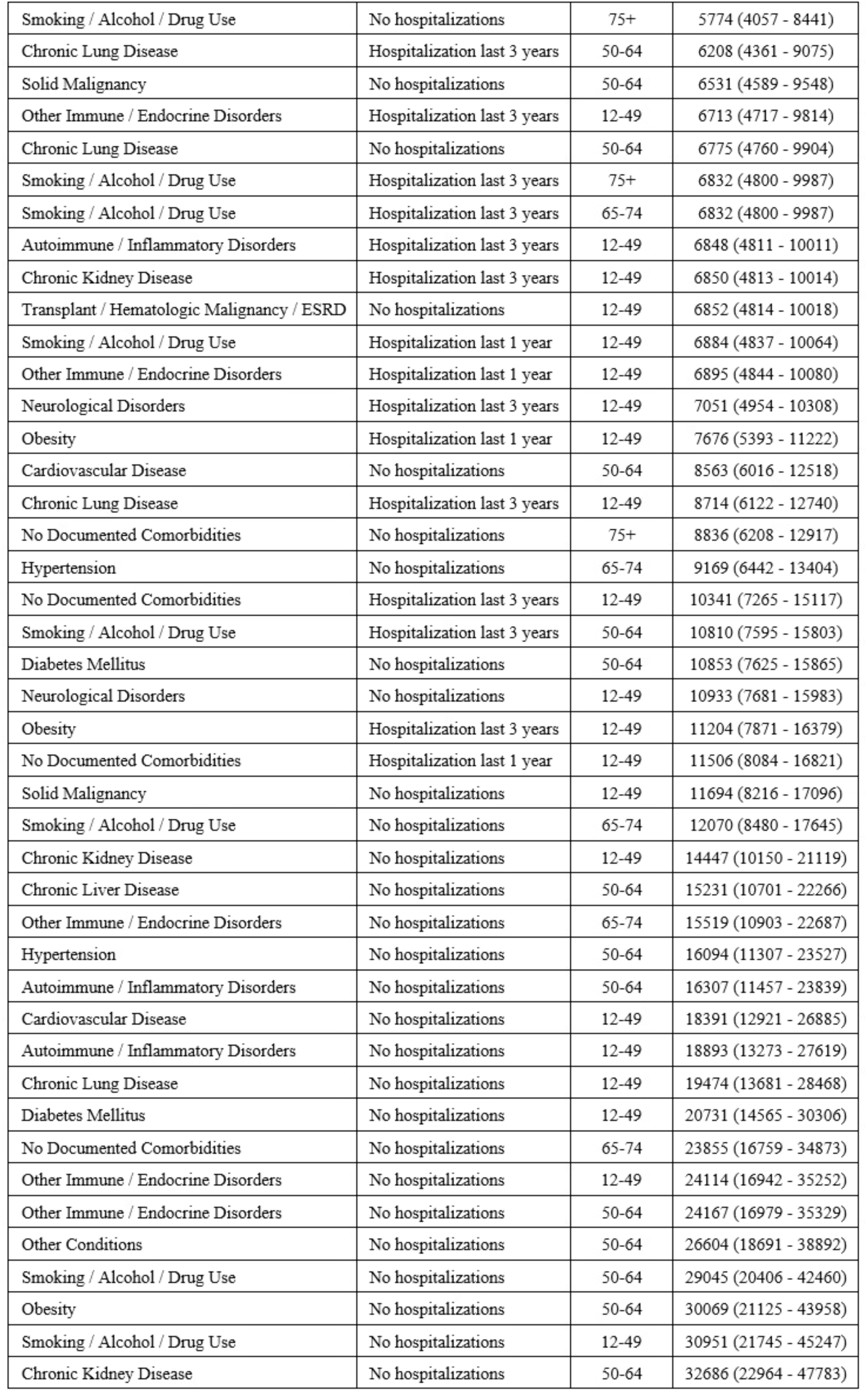
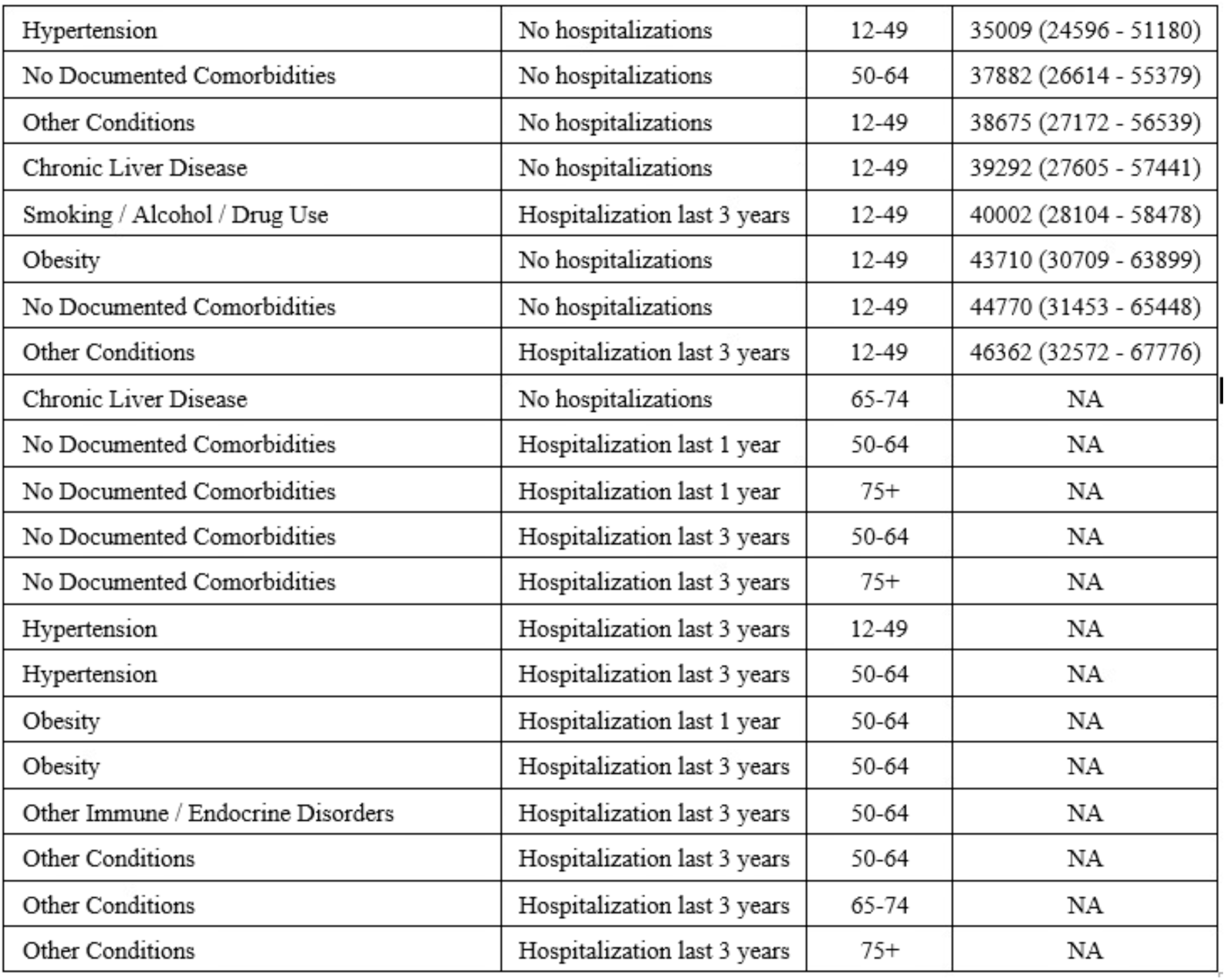
Number needed to vaccinate (NNV), with 95% CI, to prevent 1 moderate-to-severe COVID-19 hospitalization for each subgroup during the post-emergency phase.

| **Health group** | **Hospitalizations history** | **Age group** | **NNV** |
| --- | --- | --- | --- |
| Transplant / Hematologic Malignancy / ESRD | Hospitalization last 1 year | 75+ | 69 (48 - 100) |
| Transplant / Hematologic Malignancy / ESRD | Hospitalization last 1 year | 65-74 | 89 (63 - 131) |
| Severe Pulmonary or Hepatic Disease | Hospitalization last 1 year | 75+ | 91 (64 - 133) |
| Transplant / Hematologic Malignancy / ESRD | Hospitalization last 1 year | 50-64 | 147 (103 - 214) |
| Severe Pulmonary or Hepatic Disease | Hospitalization last 1 year | 65-74 | 166 (116 - 242) |
| Transplant / Hematologic Malignancy / ESRD | Hospitalization last 3 years | 75+ | 174 (122 - 254) |
| Solid Malignancy | Hospitalization last 1 year | 75+ | 178 (125 - 260) |
| Cardiovascular Disease | Hospitalization last 1 year | 75+ | 186 (131 - 272) |
| Severe Pulmonary or Hepatic Disease | Hospitalization last 3 years | 75+ | 194 (137 - 284) |
| Severe Pulmonary or Hepatic Disease | Hospitalization last 1 year | 50-64 | 211 (148 - 308) |
| Transplant / Hematologic Malignancy / ESRD | Hospitalization last 1 year | 12-49 | 261 (183 - 381) |
| Neurological Disorders | No hospitalizations | 75+ | 267 (188 - 391) |
| Severe Pulmonary or Hepatic Disease | No hospitalizations | 75+ | 280 (197 - 410) |
| Chronic Kidney Disease | Hospitalization last 1 year | 75+ | 302 (212 - 441) |
| Transplant / Hematologic Malignancy / ESRD | Hospitalization last 3 years | 65-74 | 307 (215 - 448) |
| Transplant / Hematologic Malignancy / ESRD | No hospitalizations | 75+ | 319 (224 - 466) |
| Autoimmune / Inflammatory Disorders | Hospitalization last 1 year | 75+ | 324 (228 - 474) |
| Solid Malignancy | Hospitalization last 3 years | 75+ | 337 (236 - 492) |
| Cardiovascular Disease | Hospitalization last 3 years | 75+ | 344 (242 - 503) |
| Severe Pulmonary or Hepatic Disease | Hospitalization last 1 year | 12-49 | 358 (252 - 524) |
| Neurological Disorders | Hospitalization last 1 year | 75+ | 364 (256 - 532) |
| Neurological Disorders | Hospitalization last 1 year | 65-74 | 364 (256 - 532) |
| Solid Malignancy | Hospitalization last 1 year | 65-74 | 371 (260 - 542) |
| Hypertension | Hospitalization last 1 year | 65-74 | 379 (266 - 554) |
| Hypertension | Hospitalization last 1 year | 75+ | 379 (266 - 554) |
| Transplant / Hematologic Malignancy / ESRD | Hospitalization last 3 years | 50-64 | 430 (302 - 629) |
| Severe Pulmonary or Hepatic Disease | Hospitalization last 3 years | 65-74 | 464 (326 - 679) |
| Solid Malignancy | No hospitalizations | 75+ | 529 (371 - 773) |
| Obesity | Hospitalization last 3 years | 75+ | 544 (382 - 796) |
| Obesity | Hospitalization last 3 years | 65-74 | 544 (382 - 796) |
| Chronic Kidney Disease | No hospitalizations | 75+ | 552 (388 - 807) |
| Solid Malignancy | Hospitalization last 1 year | 50-64 | 575 (404 - 840) |
| Cardiovascular Disease | Hospitalization last 1 year | 65-74 | 577 (405 - 843) |
| Cardiovascular Disease | No hospitalizations | 75+ | 595 (418 - 870) |
| Diabetes Mellitus | No hospitalizations | 75+ | 597 (419 - 872) |
| Transplant / Hematologic Malignancy / ESRD | No hospitalizations | 65-74 | 633 (444 - 925) |
| Chronic Lung Disease | Hospitalization last 3 years | 65-74 | 651 (457 - 951) |
| Chronic Lung Disease | Hospitalization last 3 years | 75+ | 651 (457 - 951) |
| Chronic Lung Disease | Hospitalization last 1 year | 50-64 | 677 (476 - 989) |
| Severe Pulmonary or Hepatic Disease | Hospitalization last 3 years | 50-64 | 716 (503 - 1046) |
| Chronic Lung Disease | Hospitalization last 1 year | 75+ | 723 (508 - 1058) |
| Chronic Lung Disease | Hospitalization last 1 year | 65-74 | 723 (508 - 1058) |
| Chronic Lung Disease | No hospitalizations | 75+ | 744 (522 - 1087) |
| Severe Pulmonary or Hepatic Disease | No hospitalizations | 65-74 | 790 (555 - 1155) |
| Diabetes Mellitus | Hospitalization last 1 year | 50-64 | 801 (563 - 1171) |
| Diabetes Mellitus | Hospitalization last 1 year | 75+ | 814 (572 - 1191) |
| Diabetes Mellitus | Hospitalization last 1 year | 65-74 | 814 (572 - 1191) |
| Chronic Kidney Disease | Hospitalization last 1 year | 65-74 | 820 (576 - 1199) |
| Diabetes Mellitus | Hospitalization last 3 years | 65-74 | 878 (617 - 1284) |
| Diabetes Mellitus | Hospitalization last 3 years | 75+ | 878 (617 - 1284) |
| Hypertension | No hospitalizations | 75+ | 960 (674 - 1403) |
| Chronic Liver Disease | Hospitalization last 1 year | 50-64 | 961 (675 - 1405) |
| Chronic Liver Disease | Hospitalization last 1 year | 65-74 | 975 (685 - 1425) |
| Chronic Liver Disease | Hospitalization last 1 year | 75+ | 975 (685 - 1425) |
| Chronic Liver Disease | Hospitalization last 1 year | 12-49 | 991 (697 - 1449) |
| Diabetes Mellitus | Hospitalization last 3 years | 50-64 | 1032 (725 - 1509) |
| Chronic Kidney Disease | Hospitalization last 3 years | 75+ | 1033 (726 - 1511) |
| Cardiovascular Disease | Hospitalization last 1 year | 50-64 | 1074 (754 - 1570) |
| Neurological Disorders | Hospitalization last 3 years | 75+ | 1133 (796 - 1656) |
| Neurological Disorders | Hospitalization last 3 years | 65-74 | 1133 (796 - 1656) |
| Chronic Kidney Disease | Hospitalization last 1 year | 50-64 | 1190 (836 - 1739) |
| Smoking / Alcohol / Drug Use | Hospitalization last 1 year | 75+ | 1195 (840 - 1747) |
| Smoking / Alcohol / Drug Use | Hospitalization last 1 year | 65-74 | 1195 (840 - 1747) |
| Neurological Disorders | No hospitalizations | 65-74 | 1227 (862 - 1794) |
| Severe Pulmonary or Hepatic Disease | Hospitalization last 3 years | 12-49 | 1232 (865 - 1801) |
| Autoimmune / Inflammatory Disorders | Hospitalization last 3 years | 75+ | 1250 (878 - 1827) |
| Cardiovascular Disease | Hospitalization last 3 years | 65-74 | 1274 (895 - 1863) |
| Solid Malignancy | Hospitalization last 1 year | 12-49 | 1289 (906 - 1885) |
| Autoimmune / Inflammatory Disorders | Hospitalization last 3 years | 65-74 | 1308 (919 - 1912) |
| Autoimmune / Inflammatory Disorders | No hospitalizations | 75+ | 1325 (931 - 1937) |
| Autoimmune / Inflammatory Disorders | Hospitalization last 1 year | 65-74 | 1330 (934 - 1944) |
| Other Immune / Endocrine Disorders | Hospitalization last 1 year | 65-74 | 1389 (976 - 2031) |
| Other Immune / Endocrine Disorders | Hospitalization last 1 year | 75+ | 1389 (976 - 2031) |
| Solid Malignancy | Hospitalization last 3 years | 65-74 | 1401 (984 - 2047) |
| Other Immune / Endocrine Disorders | Hospitalization last 1 year | 50-64 | 1478 (1038 - 2161) |
| Hypertension | Hospitalization last 3 years | 75+ | 1509 (1060 - 2205) |
| Hypertension | Hospitalization last 3 years | 65-74 | 1509 (1060 - 2205) |
| Obesity | Hospitalization last 1 year | 65-74 | 1529 (1074 - 2235) |
| Obesity | Hospitalization last 1 year | 75+ | 1529 (1074 - 2235) |
| Severe Pulmonary or Hepatic Disease | No hospitalizations | 50-64 | 1730 (1215 - 2529) |
| Neurological Disorders | Hospitalization last 3 years | 50-64 | 1831 (1286 - 2677) |
| Transplant / Hematologic Malignancy / ESRD | Hospitalization last 3 years | 12-49 | 1912 (1343 - 2795) |
| Solid Malignancy | Hospitalization last 3 years | 50-64 | 1955 (1373 - 2857) |
| Hypertension | Hospitalization last 1 year | 50-64 | 2023 (1422 - 2958) |
| Chronic Kidney Disease | Hospitalization last 3 years | 50-64 | 2035 (1429 - 2974) |
| Smoking / Alcohol / Drug Use | No hospitalizations | 75+ | 5774 (4057 - 8441) |
| Chronic Lung Disease | Hospitalization last 3 years | 50-64 | 6208 (4361 - 9075) |
| Solid Malignancy | No hospitalizations | 50-64 | 6531 (4589 - 9548) |
| Other Immune / Endocrine Disorders | Hospitalization last 3 years | 12-49 | 6713 (4717 - 9814) |
| Chronic Lung Disease | No hospitalizations | 50-64 | 6775 (4760 - 9904) |
| Smoking / Alcohol / Drug Use | Hospitalization last 3 years | 75+ | 6832 (4800 - 9987) |
| Smoking / Alcohol / Drug Use | Hospitalization last 3 years | 65-74 | 6832 (4800 - 9987) |
| Autoimmune / Inflammatory Disorders | Hospitalization last 3 years | 12-49 | 6848 (4811 - 10011) |
| Chronic Kidney Disease | Hospitalization last 3 years | 12-49 | 6850 (4813 - 10014) |
| Transplant / Hematologic Malignancy / ESRD | No hospitalizations | 12-49 | 6852 (4814 - 10018) |
| Smoking / Alcohol / Drug Use | Hospitalization last 1 year | 12-49 | 6884 (4837 - 10064) |
| Other Immune / Endocrine Disorders | Hospitalization last 1 year | 12-49 | 6895 (4844 - 10080) |
| Neurological Disorders | Hospitalization last 3 years | 12-49 | 7051 (4954 - 10308) |
| Obesity | Hospitalization last 1 year | 12-49 | 7676 (5393 - 11222) |
| Cardiovascular Disease | No hospitalizations | 50-64 | 8563 (6016 - 12518) |
| Chronic Lung Disease | Hospitalization last 3 years | 12-49 | 8714 (6122 - 12740) |
| No Documented Comorbidities | No hospitalizations | 75+ | 8836 (6208 - 12917) |
| Hypertension | No hospitalizations | 65-74 | 9169 (6442 - 13404) |
| No Documented Comorbidities | Hospitalization last 3 years | 12-49 | 10341 (7265 - 15117) |
| Smoking / Alcohol / Drug Use | Hospitalization last 3 years | 50-64 | 10810 (7595 - 15803) |
| Diabetes Mellitus | No hospitalizations | 50-64 | 10853 (7625 - 15865) |
| Neurological Disorders | No hospitalizations | 12-49 | 10933 (7681 - 15983) |
| Obesity | Hospitalization last 3 years | 12-49 | 11204 (7871 - 16379) |
| No Documented Comorbidities | Hospitalization last 1 year | 12-49 | 11506 (8084 - 16821) |
| Solid Malignancy | No hospitalizations | 12-49 | 11694 (8216 - 17096) |
| Smoking / Alcohol / Drug Use | No hospitalizations | 65-74 | 12070 (8480 - 17645) |
| Chronic Kidney Disease | No hospitalizations | 12-49 | 14447 (10150 - 21119) |
| Chronic Liver Disease | No hospitalizations | 50-64 | 15231 (10701 - 22266) |
| Other Immune / Endocrine Disorders | No hospitalizations | 65-74 | 15519 (10903 - 22687) |
| Hypertension | No hospitalizations | 50-64 | 16094 (11307 - 23527) |
| Autoimmune / Inflammatory Disorders | No hospitalizations | 50-64 | 16307 (11457 - 23839) |
| Cardiovascular Disease | No hospitalizations | 12-49 | 18391 (12921 - 26885) |
| Autoimmune / Inflammatory Disorders | No hospitalizations | 12-49 | 18893 (13273 - 27619) |
| Chronic Lung Disease | No hospitalizations | 12-49 | 19474 (13681 - 28468) |
| Diabetes Mellitus | No hospitalizations | 12-49 | 20731 (14565 - 30306) |
| No Documented Comorbidities | No hospitalizations | 65-74 | 23855 (16759 - 34873) |
| Other Immune / Endocrine Disorders | No hospitalizations | 12-49 | 24114 (16942 - 35252) |
| Other Immune / Endocrine Disorders | No hospitalizations | 50-64 | 24167 (16979 - 35329) |
| Other Conditions | No hospitalizations | 50-64 | 26604 (18691 - 38892) |
| Smoking / Alcohol / Drug Use | No hospitalizations | 50-64 | 29045 (20406 - 42460) |
| Obesity | No hospitalizations | 50-64 | 30069 (21125 - 43958) |
| Smoking / Alcohol / Drug Use | No hospitalizations | 12-49 | 30951 (21745 - 45247) |
| Chronic Kidney Disease | No hospitalizations | 50-64 | 32686 (22964 - 47783) |
| Hypertension | No hospitalizations | 12-49 | 35009 (24596 - 51180) |
| No Documented Comorbidities | No hospitalizations | 50-64 | 37882 (26614 - 55379) |
| Other Conditions | No hospitalizations | 12-49 | 38675 (27172 - 56539) |
| Chronic Liver Disease | No hospitalizations | 12-49 | 39292 (27605 - 57441) |
| Smoking / Alcohol / Drug Use | Hospitalization last 3 years | 12-49 | 40002 (28104 - 58478) |
| Obesity | No hospitalizations | 12-49 | 43710 (30709 - 63899) |
| No Documented Comorbidities | No hospitalizations | 12-49 | 44770 (31453 - 65448) |
| Other Conditions | Hospitalization last 3 years | 12-49 | 46362 (32572 - 67776) |
| Chronic Liver Disease | No hospitalizations | 65-74 | NA |
| No Documented Comorbidities | Hospitalization last 1 year | 50-64 | NA |
| No Documented Comorbidities | Hospitalization last 1 year | 75+ | NA |
| No Documented Comorbidities | Hospitalization last 3 years | 50-64 | NA |
| No Documented Comorbidities | Hospitalization last 3 years | 75+ | NA |
| Hypertension | Hospitalization last 3 years | 12-49 | NA |
| Hypertension | Hospitalization last 3 years | 50-64 | NA |
| Obesity | Hospitalization last 1 year | 50-64 | NA |
| Obesity | Hospitalization last 3 years | 50-64 | NA |
| Other Immune / Endocrine Disorders | Hospitalization last 3 years | 50-64 | NA |
| Other Conditions | Hospitalization last 3 years | 50-64 | NA |
| Other Conditions | Hospitalization last 3 years | 65-74 | NA |
| Other Conditions | Hospitalization last 3 years | 75+ | NA |

**Table S5.** Logistic regression results.

| **Variable** | **Coefficient value (95% CI)** | **P value** | **OR (95% CI)** |
| --- | --- | --- | --- |
| Intercept | -13.481 (-13.482 to -13.480) | <0.001 | 0.000 (0.000 to 0.000) |
| Hospitalizations in recent 3 years | 3.150 (3.149 to 3.151) | <0.001 | 23.33 (23.31 to 23.36) |
| Hospitalizations in the year prior | 3.473 (3.472 to 3.475) | <0.001 | 32.23 (32.20 to 32.26) |
| Age group | 2.149 (2.148 to 2.150) | <0.001 | 8.58 (8.57 to 8.59) |
| Age × Hospitalizations in recent 3 years | -4.301 (-4.304 to -4.298) | <0.001 | 0.013 (0.013 to 0.014) |
| Age × Hospitalizations in the year prior | -4.511 (-4.514 to -4.508) | <0.001 | 0.011 (0.011 to 0.012) |
| Health group | 0.101 (0.101 to 0.102) | <0.001 | 1.106 (1.106 to 1.107) |
| Hospitalizations in recent 3 years × Health group | -0.086 (-0.086 to -0.085) | <0.001 | 0.918 (0.918 to 0.919) |
| Hospitalizations in the year prior × Health group | -0.078 (-0.078 to -0.077) | <0.001 | 0.925 (0.926 to 0.926) |
| Age × Health group | -0.154 (-0.154 to -0.153) | <0.001 | 0.857 (0.857 to 0.858) |
| Age × Hospitalizations in recent 3 years × Health group | 0.316 (0.316 to 0.317) | <0.001 | 1.372 (1.371 to 1.374) |
| Age × Hospitalizations in the year prior × Health group | 0.325 (0.325 to 0.326) | <0.001 | 1.385 (1.384 to 1.387) |

